# Prevalence and determinants of serum antibodies to SARS-CoV-2 in the general population of the Gardena Valley

**DOI:** 10.1101/2021.03.19.21253883

**Authors:** Roberto Melotti, Federica Scaggiante, Michela Falciani, Christian X. Weichenberger, Luisa Foco, Stefano Lombardo, Alessandro De Grandi, Dorothee von Laer, Angelika Mahlknecht, Peter P. Pramstaller, Elisabetta Pagani, Horand Meier, Timon Gaertner, Christina Troi, Deborah Mascalzoni, Cristian Pattaro, Michael Mian

## Abstract

**Background:** Community-based studies are essential to quantify the spread of SARS-CoV-2 infection and for unbiased characterization of its determinants and outcomes. We conducted a cross-sectional study in the Gardena valley, a major Alpine touristic destination which was struck in the expansion phase of the COVID-19 pandemic over the winter 2020.

**Methods:** We surveyed 2244 representative study participants who underwent swab and serum antibody tests. We made multiple comparisons among the Abbott and Diasorin bioassays and serum neutralization titers. Seroprevalence accounted for the stratified design, non-response and test accuracy. Determinants and symptoms predictive of infection were analyzed by weighted multiple logistic regression.

**Results:** SARS-CoV-2 seroprevalence was 26.9% (95% confidence interval: 25.2%, 28.6%) by June 2020. The serum antibody bioassays had modest agreement with each other. Receiver operating characteristic curve analysis on the serum neutralizing capacity showed better performance of the Abbott test at lower than the canonical threshold. Socio-demographic characteristics showed no clear evidence of association with seropositivity, which was instead associated with place of residence and economic activity. Loss of taste or smell, fever, difficulty in breathing, pain in the limbs, and weakness were the most predictive symptoms of positive antibody test results. Fever and weakness associations were age-dependent.

**Conclusion:** The Gardena valley had one of the highest SARS-CoV-2 infection prevalence in Europe. The age-dependent risk associated with COVID-19 related symptoms implies targeted strategies for screening and prophylaxis planning.

## INTRODUCTION

During the initial phase of the SARS-CoV-2 pandemic, the Gardena valley, a well-known winter tourism destination located in South Tyrol (Italy), became one of the European regions most afflicted by the coronavirus disease 2019 (COVID-19). While in the middle of the virus circulation vortex since February 2020, there were a multitude of holidaymakers and visitors in the valley mainly from Northern Italy and Central Europe. Back home, tourists likely contributed to further transmission of the virus just before containment actions were endorsed by regions worldwide.[1]

As expected in such an emergent phase of the pandemic, hospital-based case reports dominated the accumulation of scientific evidence on COVID-19.[2] Consequently, public awareness, ongoing knowledge of the determinants of disease and disease severity, and current prevention strategies have been profoundly influenced by clinical observations, while evidence from community studies has had limited space in context.[3] Specific knowledge of the exogenous determinants of SARS-CoV-2 infection and its related symptoms or about biological susceptibility in the general population is still incomplete, probably due to the slower pace and relative paucity of community-based studies.[4] Geographically confined regions with a relatively high incidence of infection may help characterize the spread of COVID-19, providing useful indications to policy-makers for current and future preventive efforts.

At the end of May 2020, we surveyed 2244 inhabitants of the Gardena valley representative of the local population, measured antibody test response to SARS-CoV-2 and related that response to symptoms, prior conditions and serum neutralizing capacity. The high seroprevalence qualified the in-depth analysis of determinants and COVID-19-related symptoms in a general population setting, augmenting the general understanding of the disease dynamic.

## MATERIALS AND METHODS

### Study design

Invited to the study were 2958 of the 9424 inhabitants of Ortisei, Santa Cristina, and Selva, the main municipalities of the Gardena valley, following a one-stage random sampling design stratified by municipality, sex and age group (<6, 6-17, 18-34, 35-49, 50-64, 65+ years). Sample size was defined based on an expected 3% minimal seroprevalence with 0.25% relative standard error (SE) and accounting for finite population correction. Participants were selected with known extraction probability from the municipality registries, excluding nursing homes, using the ‘surveyselect’ program in SAS v9.2.

Participants were invited via letter including: the planned participation date; a link to the online questionnaire (with telephone support) covering demographic, clinical and socio-behavioral aspects (Suppl. Mat. page 2); a personalized password for use as pseudo-anonymization code. Testing procedures included a nasopharyngeal swab test and a serological antibody test (limited to 6+ year old participants). The study took place between May 26 and June 8, 2020. The Ethics Committee of the Healthcare System of the Autonomous Province of Bolzano/Bozen authorized the study. Each participant gave written informed consent.

### Biological sample collection and analysis

Swab samples were analyzed at the ÖNORM-accredited (EN ISO 15189:2013) diagnostic laboratory of the Institute of Virology of the Innsbruck Medical University (IVIMU, Austria) as described in the Suppl. Mat. page 7. As no swab sample tested positive, this analysis was not further considered.

Antibody response was tested using the Abbott SARS-CoV-2 IgG assay (Sligo, Ireland), designed to detect immunoglobulin class G (IgG) antibodies to the nucleocapsid (N) protein of SARS-CoV-2. Fresh serum samples were collected in blood tubes with separating gel. Within 6 hours from collection, assessment of IgG antibodies to SARS-CoV-2 was performed using the Abbott Architect i2000SR system, which implements a two-step chemiluminescent microparticle immunoassay, at Laboratory of Clinical Pathology of the Bressanone/Brixen Hospital, Italy. Seropositivity was defined as a signal-to-calibrator (S/C) Abbott Architect Index (AAI) value ≥ 1.4. At this threshold, the manufacturer reported 96.9% (89.5% to 99.5%) sensitivity at 14 days after symptoms onset, 100.0% (95.1% to 100.0%) sensitivity at 17 days, and 99.9% specificity.[5] Biological samples of study participants were stored at the Eurac Research Biobank (ERB) at the Bolzano/Bozen Hospital, Italy, as described in the Suppl. Mat. page 7.

Two hundred ninety-nine serum samples were selected for plaque reduction neutralization test (PRNT),[6] ensuring the coverage of the whole SARS-CoV-2 IgG assay AAI distribution while maximizing sample heterogeneity in terms of sex, age, and symptoms manifestation as well as previous diagnosis and hospitalization (additional details in Suppl. Mat. page 7 and Suppl. Fig. 1A and 1B). Selected frozen serum samples were dry ice shipped from the ERB to the IVIMU diagnostic laboratory. After 30 minute heat inactivation at 56°C, samples were tabletop centrifuged for 5 minutes at 8000 rpm. They were 4-fold serially diluted in complete medium containing 2% fetal calf serum (FCS) starting with a 1:4 dilution in duplicate samples. Serum dilutions were mixed with an equal volume of a replication competent SARS-CoV-2 resulting in ∼300 infected cells in non-neutralized wells. Serum/virus mixes were incubated for 1 hour at 37°C and subsequently transferred to 96-wells containing 90% confluent Vero cells expressing TMPRSS2 seeded one day before. Cells were infected with the virus for 1 hour at 37°C and subsequently washed once with complete medium with 2% FCS. After adding fresh complete medium containing 2% FCS, cells were further cultured for 13 hours. Cells were fixed for 5 minutes with 96% ethanol and subsequently stained using the serum from a SARS-CoV-2 recovered patient and a horse radish peroxidase-conjugated anti-human secondary antibody (Dianova). Plates were developed using 3-amino-9-ethylcarbazole substrate. Infected cells were counted via microscope and 50% neutralization titers were calculated as the highest dilution where the mean infection of duplicate samples was reduced by >50% of the mean of control wells lacking the virus. The presence of spike (S) protein antibodies in the 299 samples was assessed with the Diasorin LIAISON® SARS-CoV-2 S1/S2 IgG chemiluminescent assay (Saluggia, Italy), designed to detect the number of arbitrary units (AUs) of specific IgG class antibodies directed against the S1 and S2 viral proteins, at the Microbiology and Virology laboratory of the South Tyrolean Healthcare System (Bolzano/Bozen, Italy). Results were classified as negative (values<12 AU/ml), inconclusive (values≥ 12 and <15 AU/ml), or positive (values≥ 15 AU/ml) according to manufacturer’s recommendations. This assay had declared 97.4% sensitivity >15 days after symptoms onset, and 98.9% specificity.[7]

### Statistical analyses

We assessed pairwise agreement between quantitative variables with the Lin’s concordance correlation coefficient (CCC)[8] and Bland-Altman plot,[9] and agreement between categorical variables using the Cohen’s kappa statistic,[10] using the ‘epiR’ v1.0-15, ‘BlandAltmanLeh’ v0.3.1, and ‘psych’ v1.9.11 packages in the R software v3.3.6.

To investigate the discrimination accuracy of the 1.4 AAI threshold on thawed serum against a PRNT value≥ 4, which was considered as a gold-standard for prior exposure to SARS-CoV-2, we conducted receiver operating characteristic (ROC) curve analysis identifying the optimal cutoff as the Youden’s index (*J*).[11] Overfitting was prevented by performing both 10-fold cross-validation and repeated random sub-sampling validation. In the latter approach, we randomly split the sample set into 80% and 20% training and test sets, respectively, corresponding to 239 and 60 observations, repeatedly 5000 times. The two validation procedures resulted in sets of 10 and 5000 optimal cutoffs, respectively. We reported the median optimal cutoff of each respective validation procedure.

Descriptive tables and multiple logistic regression models display observed counts for each relevant category, while accounting for the study design: stratification by sex, municipality and age group, post-stratification (citizenship by municipality) and finite population correction, for efficient proportion estimations. To correct for possible selection biases, we also adjusted the sampling weights based on proportional allocation within strata by non-response (balanced to the population strata distribution), which were then calibrated dynamically by post-stratification in each analysis. Prevalence of serum antibodies to SARS-CoV-2 (seroprevalence) in the overall sample was also estimated using the Rogan and Gladen formula to account for the serological test inaccuracy.[12] Statistical analyses were run with the ‘svyset’ suite of commands in Stata software v16.1 (StataCorp LLC) and were restricted to 6+ year old individuals and non-pregnant women using the ‘subpop’ option.

## RESULTS

Of 2958 invited individuals, 2244 (75.9%) joined the study. Among these, swab and serum antibody test results were available for 2083 (82.8%) and 2129 (94.9%) participants, respectively. Excluding pregnant women and <6 year old children left 2106 (93.9%) participants for seroprevalence analysis, of whom 1813 (80.8%) filled in the questionnaire-based interview. Participants were balanced across sexes and age groups (Table 1). In terms of sex, age, and municipality distribution, the sample was representative of the reference population. Of the 2106 participants undergoing the serum antibody test, 551 tested positive, corresponding to a corrected seroprevalence of 26.9% (95% confidence interval, 95%CI: 25.2%, 28.6%), with no evidence of different seroprevalence between questionnaire completers and non-completers (Suppl. Tab. 1).

**Table 1.**
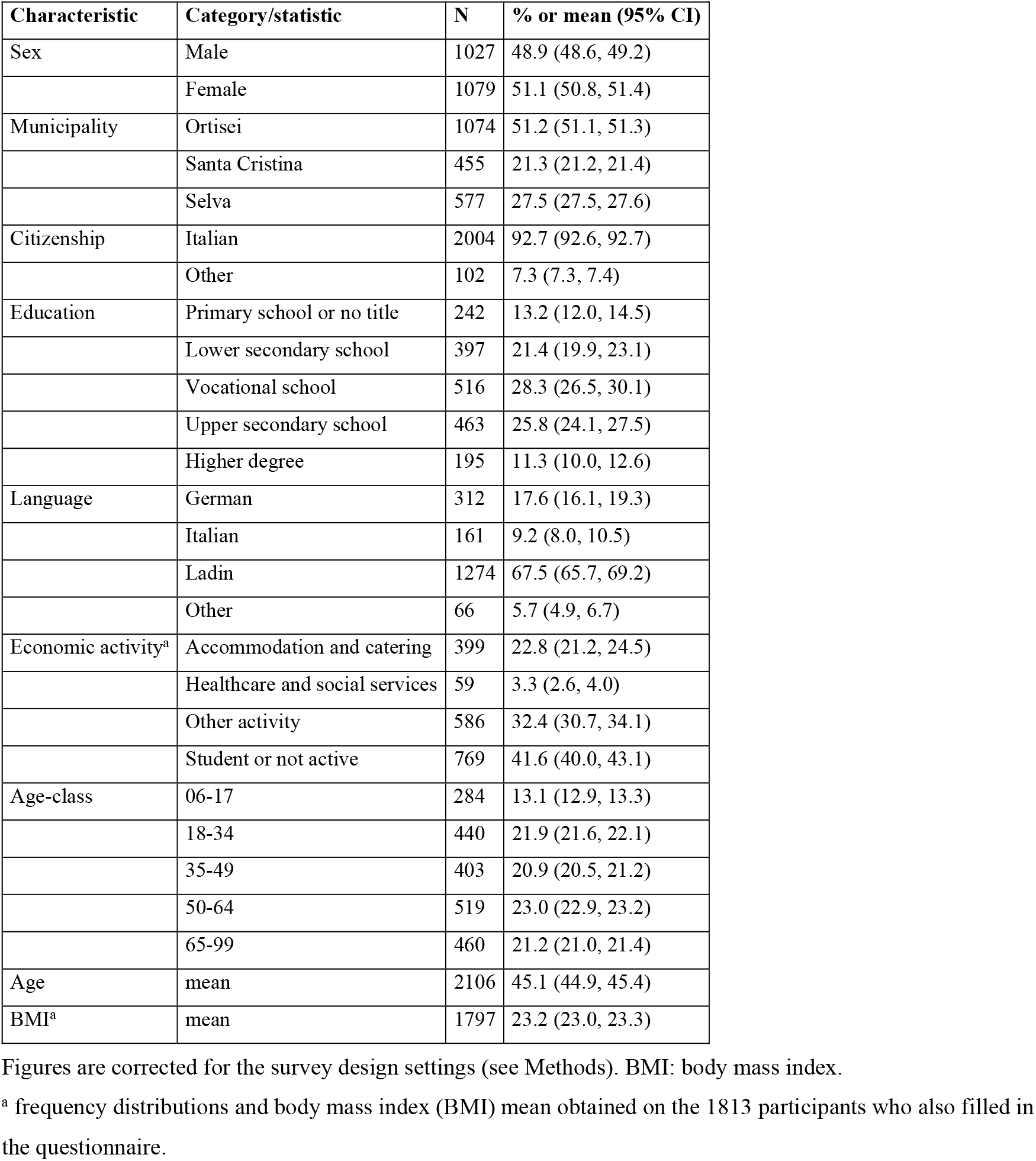
Socio-demographic characteristics of the 2106 participants with available antibody test results who were considered for the seroprevalence analysis (age 6+; non-pregnant women).

### Comparison of testing methods and plaque reduction neutralization test

The 299 participants samples chosen for PRNT analysis had similar characteristics to the whole sample (Suppl. Tab. 2). Their S/C (AAI) values were uniformly distributed across the whole range observed (Suppl. Fig. 1C), resulting in 194 seropositive samples. To exclude sample logistic, handling and thawing procedure effects, we first compared Abbott antibody test results in fresh against post-conservation thawed serum, observing almost perfect agreement (CCC=0.9982; 95%CI: 0.9978, 0.9986; Suppl. Fig. 2A) despite minimal discordance attributable to storage time (Suppl. Fig. 2B) and plate (Suppl. Fig. 2C) effects. Post-thawing sample levels were on average −0.08 (95%CI: - 0.09, −0.06) AAI units lower than those observed in fresh blood (Suppl. Fig. 2B), with two positive samples reclassified as negative (kappa=0.99; 95%CI: 0.97, 1.00). We observed 192 (64.2%) positive samples by the Abbott assay at the 1.4 canonical threshold, and 190 (63.5%) and 197 (65.9%) positive samples by the Diasorin test at the 15 and 12 thresholds, respectively (Suppl. Tab. 2, Fig. 1A). In either case, the concordance between Abbott and Diasorin assays was limited (Fig. 1A and 1B).

**Figure 1.**
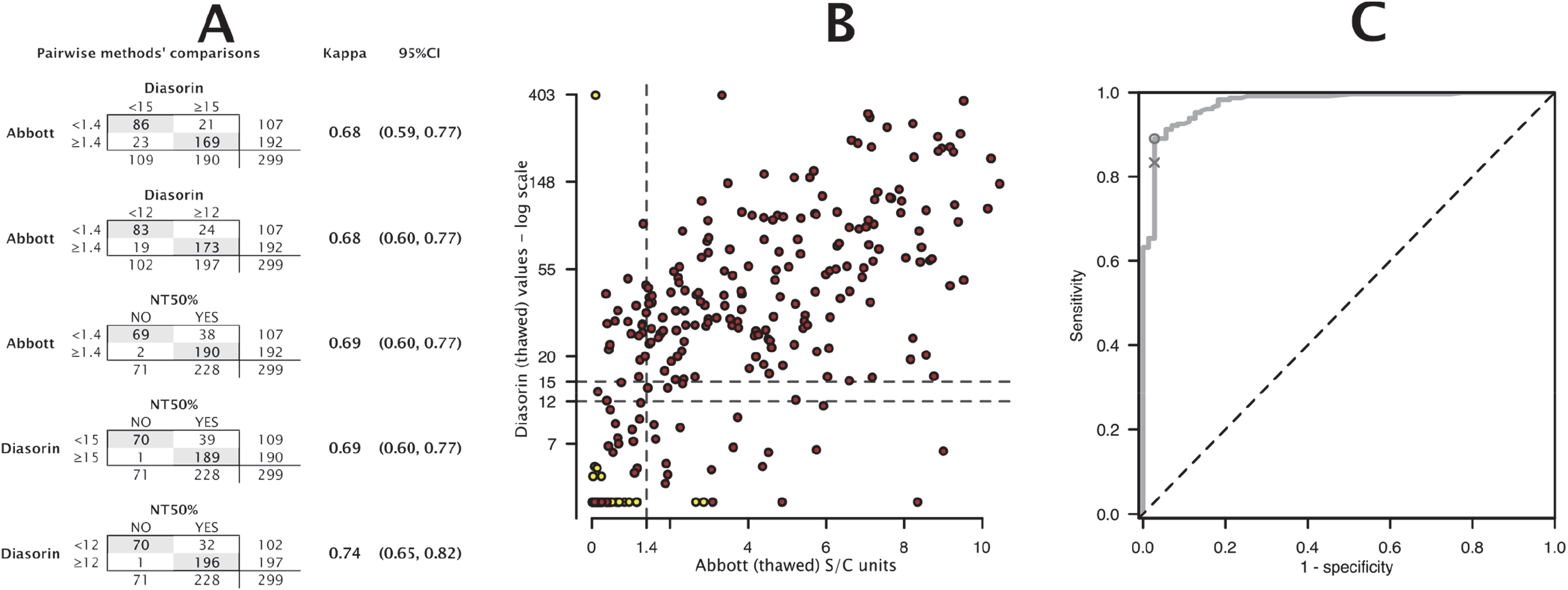
Antibody test performance evaluations. **Panel A**: Pairwise comparisons and kappa statistics with their 95%CIs across all antibody assays and PRNT at 50%. **Panel B**: Scatterplot of the Abbott assay results (x-axis) versus the Diasorin assay results (y-axis, in logarithmic scale), in the context of PRNT results (yellow dots: negative; brown dots: positive). Dashed lines indicate the clinically relevant thresholds for positivity. **Panel C**: ROC curve used to define the optimal cut-off for S/C (AAI) values as a classifier. Plotted here is the true positive rate (sensitivity) versus the false positive rate (1 – specificity) for increasing values of AAI for all 299 individuals subject to PRNT. The diagonal corresponds to the ROC curve of a random classifier. Discriminative classifiers produce curves drawn towards the upper left corner, where sensitivity = specificity = 1. A perfectly discriminating classifier generates a ROC curve that starts at the lower left corner and advances as a vertical line to the upper left corner and from there horizontally to the upper right corner. The cross corresponds to the classifier performance using AAI = 1.4. In the sample under investigation, optimal classifier performance is achieved for AAI = 1.16 (circle).

Of the 299 samples, 228 (76.3%) showed 50% neutralization capacity. This was in limited agreement with both Abbott (kappa=0.69; 95%CI: 0.60, 0.77) and Diasorin assay when evaluated at the canonical threshold (kappa=0.69; 95%CI: 0.60, 0.77; Fig. 1A). Higher agreement was observed between PRNT and Diasorin test when the latter was evaluated at a threshold of 12 (kappa=0.74; 95%CI: 0.65, 0.82). In any case, of the 83 samples testing negative with both Abbott (<1.4) and Diasorin (<12) assays, 15 (18.1%) showed neutralization capacity. In contrast, neutralization capacity was always confirmed when both the Abbott and Diasorin tests were positive, and almost always when at least one of them was positive (Fig. 1B).

The optimal Abbott antibody test threshold identified in the ROC curve analysis was 1.16 (Fig. 1C), consistently in both the 10-fold cross-validation and 5000 repeated random sub-sampling validation, both returning 1.16 median AAI. At this threshold, the classifier performed with 89.0% sensitivity and 97.2% specificity. At the recommended 1.4 threshold, sensitivity was lower (83.3%) but specificity was the same. Sex and age stratification improved the assay performance with relatively lower thresholds but at the cost of reliability (Suppl. Fig. 3; Suppl. Tab. 3).

### Identification of determinants and predictors of seropositivity

Among all sociodemographic and lifestyle characteristics considered (Table 1), being a current smoker (OR 0.35; 95%CI: 0.25, 0.49), working in the accommodation and catering services (OR=1.37; 95%CI: 1.05, 1.77), living in Selva (OR=1.37; 95%CI: 1.10, 1.70) or Santa Cristina (OR=1.31; 95%CI: 1.03, 1.68), and having higher BMI (OR=1.03; 95%CI: 1.00, 1.06) were each individually and independently associated with positivity to the Abbott antibody test (Table 2). Females did appear at lower risk than males, however, evidence was weak in the mutually adjusted analysis.

**Table 2.**
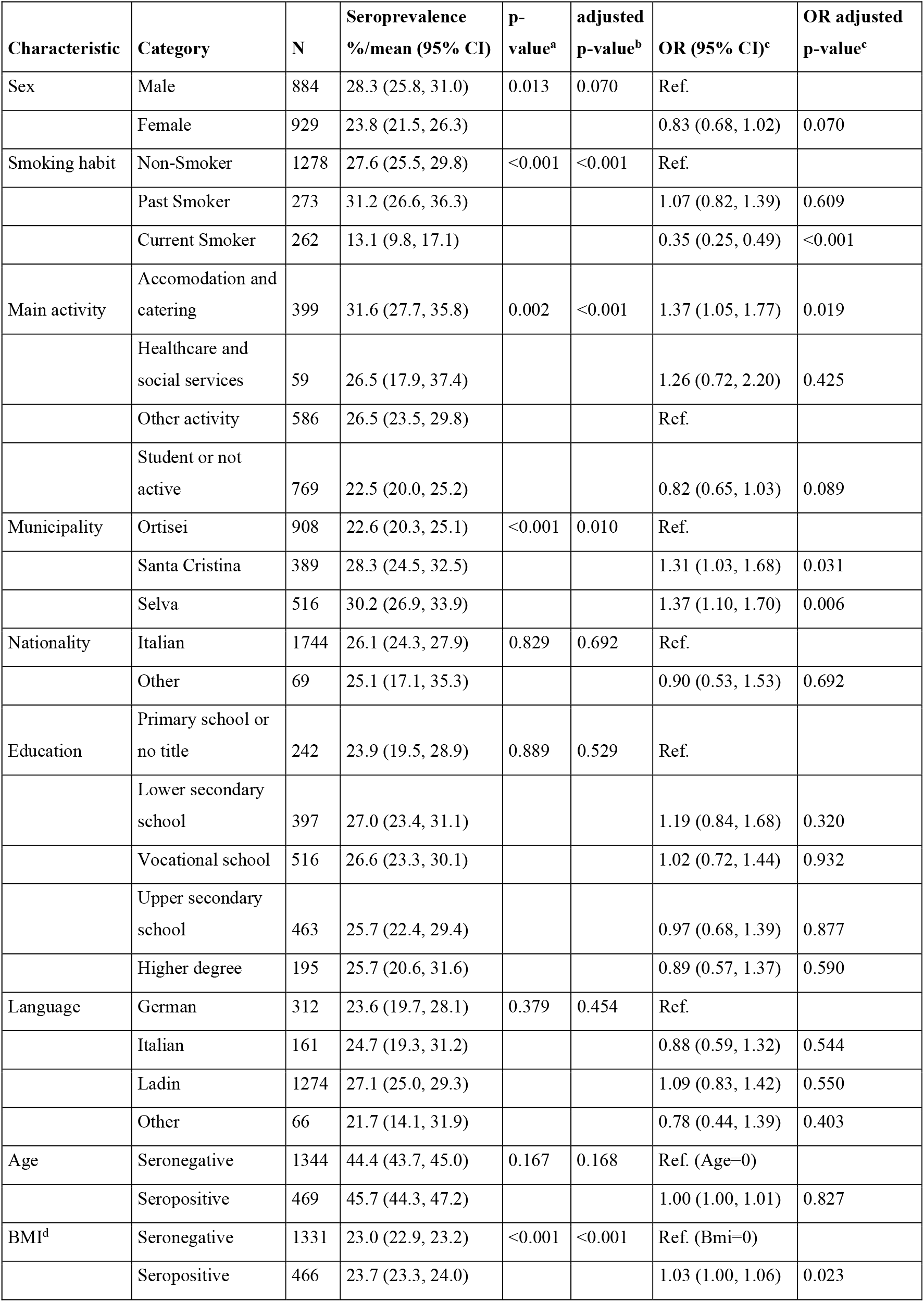
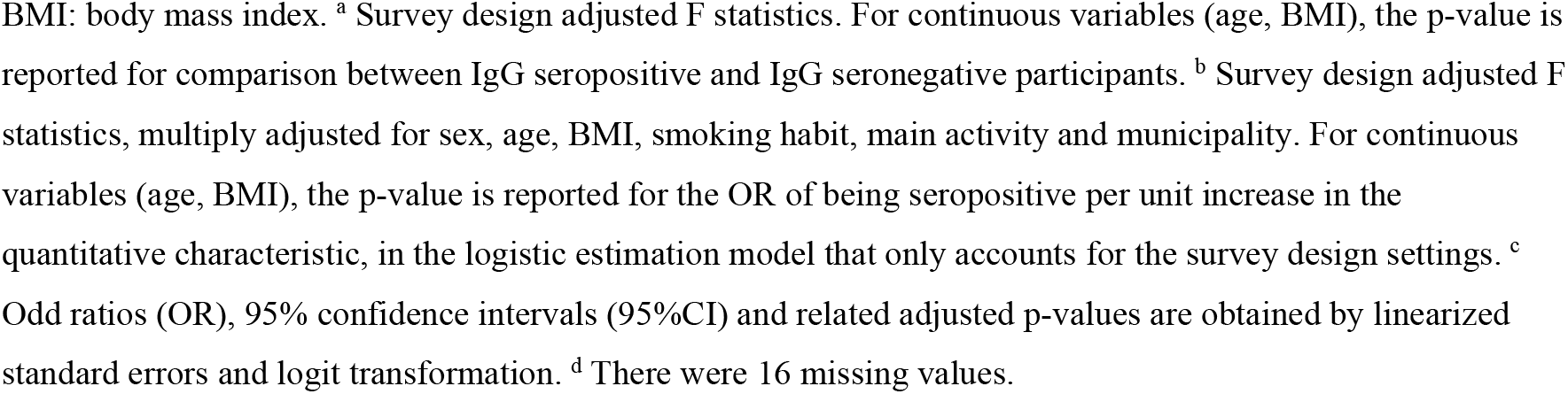
Prevalence of serum antibodies to SARS-CoV-2 and participant’s socio-demographic characteristics (N=1813).

| Characteristic | Category | N | Seroprevalence<br>%/mean (95% CI) | p-value<sup>a</sup> | adjusted<br>p-value<sup>b</sup> | OR (95% CI)<sup>c</sup> | OR adjusted<br>p-value<sup>c</sup> |
| --- | --- | --- | --- | --- | --- | --- | --- |
| Sex | Male | 884 | 28.3 (25.8, 31.0) | 0.013 | 0.070 | Ref. |  |
|  | Female | 929 | 23.8 (21.5, 26.3) |  |  | 0.83 (0.68, 1.02) | 0.070 |
| Smoking habit | Non-Smoker | 1278 | 27.6 (25.5, 29.8) | <0.001 | <0.001 | Ref. |  |
|  | Past Smoker | 273 | 31.2 (26.6, 36.3) |  |  | 1.07 (0.82, 1.39) | 0.609 |
|  | Current Smoker | 262 | 13.1 (9.8, 17.1) |  |  | 0.35 (0.25, 0.49) | <0.001 |
| Main activity | Accommodation and catering | 399 | 31.6 (27.7, 35.8) | 0.002 | <0.001 | 1.37 (1.05, 1.77) | 0.019 |
|  | Healthcare and social services | 59 | 26.5 (17.9, 37.4) |  |  | 1.26 (0.72, 2.20) | 0.425 |
|  | Other activity | 586 | 26.5 (23.5, 29.8) |  |  | Ref. |  |
|  | Student or not active | 769 | 22.5 (20.0, 25.2) |  |  | 0.82 (0.65, 1.03) | 0.089 |
| Municipality | Ortisei | 908 | 22.6 (20.3, 25.1) | <0.001 | 0.010 | Ref. |  |
|  | Santa Cristina | 389 | 28.3 (24.5, 32.5) |  |  | 1.31 (1.03, 1.68) | 0.031 |
|  | Selva | 516 | 30.2 (26.9, 33.9) |  |  | 1.37 (1.10, 1.70) | 0.006 |
| Nationality | Italian | 1744 | 26.1 (24.3, 27.9) | 0.829 | 0.692 | Ref. |  |
|  | Other | 69 | 25.1 (17.1, 35.3) |  |  | 0.90 (0.53, 1.53) | 0.692 |
| Education | Primary school or no title | 242 | 23.9 (19.5, 28.9) | 0.889 | 0.529 | Ref. |  |
|  | Lower secondary school | 397 | 27.0 (23.4, 31.1) |  |  | 1.19 (0.84, 1.68) | 0.320 |
|  | Vocational school | 516 | 26.6 (23.3, 30.1) |  |  | 1.02 (0.72, 1.44) | 0.932 |
|  | Upper secondary school | 463 | 25.7 (22.4, 29.4) |  |  | 0.97 (0.68, 1.39) | 0.877 |
|  | Higher degree | 195 | 25.7 (20.6, 31.6) |  |  | 0.89 (0.57, 1.37) | 0.590 |
| Language | German | 312 | 23.6 (19.7, 28.1) | 0.379 | 0.454 | Ref. |  |
|  | Italian | 161 | 24.7 (19.3, 31.2) |  |  | 0.88 (0.59, 1.32) | 0.544 |
|  | Ladin | 1274 | 27.1 (25.0, 29.3) |  |  | 1.09 (0.83, 1.42) | 0.550 |
|  | Other | 66 | 21.7 (14.1, 31.9) |  |  | 0.78 (0.44, 1.39) | 0.403 |
| Age | Seronegative | 1344 | 44.4 (43.7, 45.0) | 0.167 | 0.168 | Ref. (Age=0) |  |
|  | Seropositive | 469 | 45.7 (44.3, 47.2) |  |  | 1.00 (1.00, 1.01) | 0.827 |
| BMI<sup>d</sup> | Seronegative | 1331 | 23.0 (22.9, 23.2) | <0.001 | <0.001 | Ref. (Bmi=0) |  |
|  | Seropositive | 466 | 23.7 (23.3, 24.0) |  |  | 1.03 (1.00, 1.06) | 0.023 |

BMI: body mass index. <sup>a</sup> Survey design adjusted F statistics. For continuous variables (age, BMI), the p-value is reported for comparison between IgG seropositive and IgG seronegative participants. <sup>b</sup> Survey design adjusted F statistics, multiply adjusted for sex, age, BMI, smoking habit, main activity and municipality. For continuous variables (age, BMI), the p-value is reported for the OR of being seropositive per unit increase in the quantitative characteristic, in the logistic estimation model that only accounts for the survey design settings. <sup>c</sup> Odd ratios (OR), 95% confidence intervals (95%CI) and related adjusted p-values are obtained by linearized standard errors and logit transformation. <sup>d</sup> There were 16 missing values.

Focusing on symptoms as possible seroprevalence predictors, we found strong evidence for any single symptom to predict antibody positivity, both in unadjusted and mutually adjusted analyses, which considered each symptom at one time (Table 3). Seropositivity was 38.8% (95%CI: 36.2, 41.5) in those reporting any number of symptoms, 45.6% (95%CI: 42.3, 48.9) in those reporting multiple symptoms, 10.0% (95%CI: 8.3, 12.0) in those reporting no symptoms (p<0.001, Table 3), and 14.2% (95%CI: 12.4, 16.1) in those with at most 1 symptom, respectively (Table 3, Table 4).

**Table 3.**
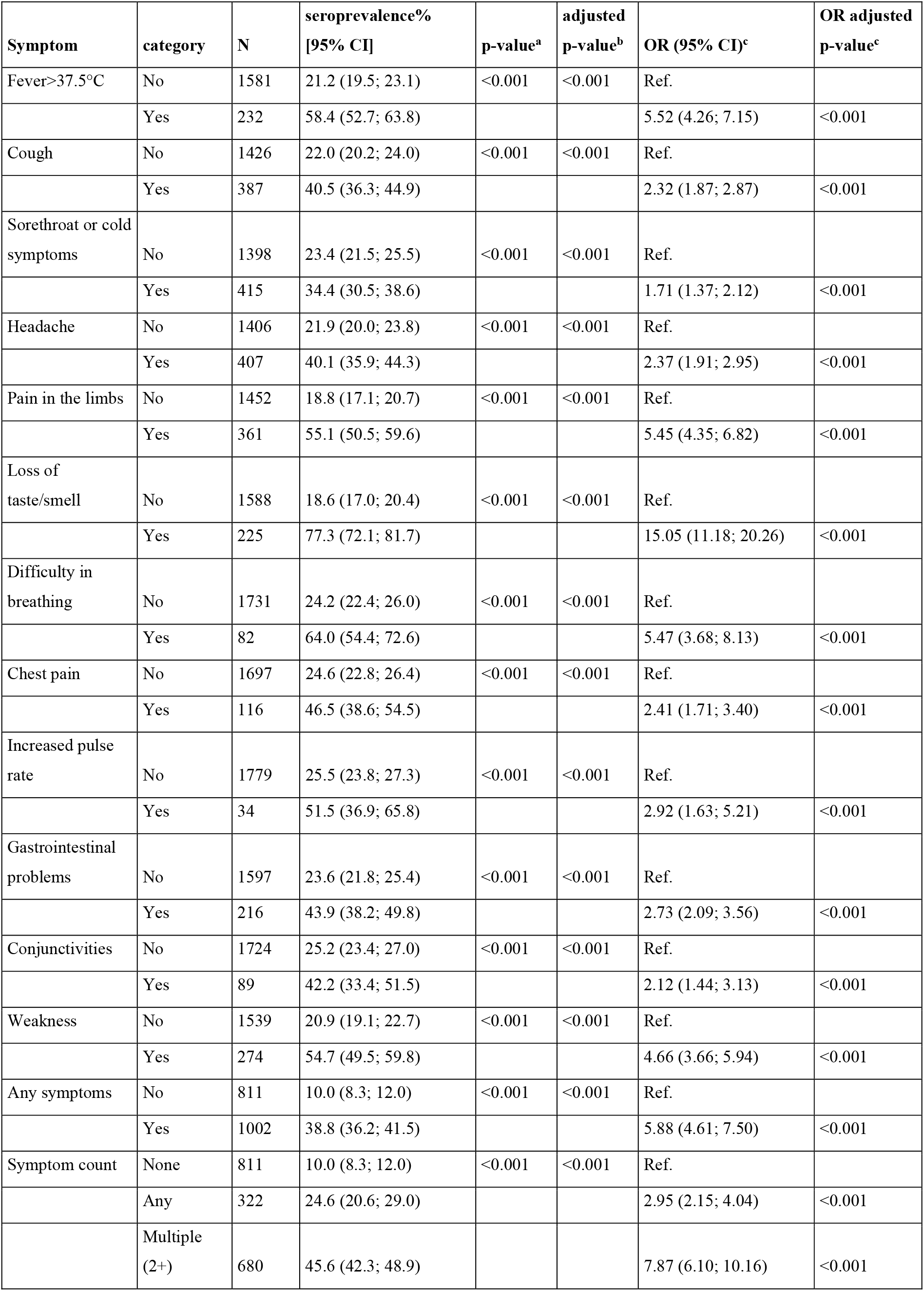
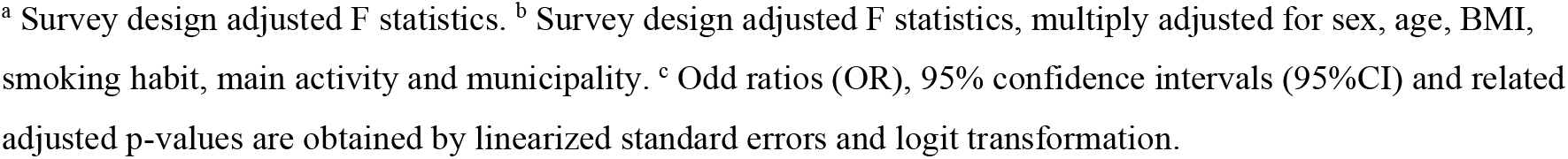
Prevalence of serum antibodies to SARS-CoV-2 by participant’s reported symptoms (N=1813).

| Symptom | category | N | seroprevalence%<br>[95% CI] | p-value<sup>a</sup> | adjusted<br>p-value<sup>b</sup> | OR (95% CI)<sup>c</sup> | OR adjusted<br>p-value<sup>c</sup> |
| --- | --- | --- | --- | --- | --- | --- | --- |
| Fever>37.5°C | No | 1581 | 21.2 (19.5; 23.1) | <0.001 | <0.001 | Ref. |  |
|  | Yes | 232 | 58.4 (52.7; 63.8) |  |  | 5.52 (4.26; 7.15) | <0.001 |
| Cough | No | 1426 | 22.0 (20.2; 24.0) | <0.001 | <0.001 | Ref. |  |
|  | Yes | 387 | 40.5 (36.3; 44.9) |  |  | 2.32 (1.87; 2.87) | <0.001 |
| Sorethroat or cold symptoms | No | 1398 | 23.4 (21.5; 25.5) | <0.001 | <0.001 | Ref. |  |
|  | Yes | 415 | 34.4 (30.5; 38.6) |  |  | 1.71 (1.37; 2.12) | <0.001 |
| Headache | No | 1406 | 21.9 (20.0; 23.8) | <0.001 | <0.001 | Ref. |  |
|  | Yes | 407 | 40.1 (35.9; 44.3) |  |  | 2.37 (1.91; 2.95) | <0.001 |
| Pain in the limbs | No | 1452 | 18.8 (17.1; 20.7) | <0.001 | <0.001 | Ref. |  |
|  | Yes | 361 | 55.1 (50.5; 59.6) |  |  | 5.45 (4.35; 6.82) | <0.001 |
| Loss of taste/smell | No | 1588 | 18.6 (17.0; 20.4) | <0.001 | <0.001 | Ref. |  |
|  | Yes | 225 | 77.3 (72.1; 81.7) |  |  | 15.05 (11.18; 20.26) | <0.001 |
| Difficulty in breathing | No | 1731 | 24.2 (22.4; 26.0) | <0.001 | <0.001 | Ref. |  |
|  | Yes | 82 | 64.0 (54.4; 72.6) |  |  | 5.47 (3.68; 8.13) | <0.001 |
| Chest pain | No | 1697 | 24.6 (22.8; 26.4) | <0.001 | <0.001 | Ref. |  |
|  | Yes | 116 | 46.5 (38.6; 54.5) |  |  | 2.41 (1.71; 3.40) | <0.001 |
| Increased pulse rate | No | 1779 | 25.5 (23.8; 27.3) | <0.001 | <0.001 | Ref. |  |
|  | Yes | 34 | 51.5 (36.9; 65.8) |  |  | 2.92 (1.63; 5.21) | <0.001 |
| Gastrointestinal problems | No | 1597 | 23.6 (21.8; 25.4) | <0.001 | <0.001 | Ref. |  |
|  | Yes | 216 | 43.9 (38.2; 49.8) |  |  | 2.73 (2.09; 3.56) | <0.001 |
| Conjunctivities | No | 1724 | 25.2 (23.4; 27.0) | <0.001 | <0.001 | Ref. |  |
|  | Yes | 89 | 42.2 (33.4; 51.5) |  |  | 2.12 (1.44; 3.13) | <0.001 |
| Weakness | No | 1539 | 20.9 (19.1; 22.7) | <0.001 | <0.001 | Ref. |  |
|  | Yes | 274 | 54.7 (49.5; 59.8) |  |  | 4.66 (3.66; 5.94) | <0.001 |
| Any symptoms | No | 811 | 10.0 (8.3; 12.0) | <0.001 | <0.001 | Ref. |  |
|  | Yes | 1002 | 38.8 (36.2; 41.5) |  |  | 5.88 (4.61; 7.50) | <0.001 |
| Symptom count | None | 811 | 10.0 (8.3; 12.0) | <0.001 | <0.001 | Ref. |  |
|  | Any | 322 | 24.6 (20.6; 29.0) |  |  | 2.95 (2.15; 4.04) | <0.001 |
|  | Multiple (2+) | 680 | 45.6 (42.3; 48.9) |  |  | 7.87 (6.10; 10.16) | <0.001 |

<sup>a</sup> Survey design adjusted F statistics. <sup>b</sup> Survey design adjusted F statistics, multiply adjusted for sex, age, BMI, smoking habit, main activity and municipality. <sup>c</sup> Odd ratios (OR), 95% confidence intervals (95%CI) and related adjusted p-values are obtained by linearized standard errors and logit transformation.

**Table 4.** Prevalence of serum antibodies to SARS-CoV-2 by participant’s reported period of symptoms (N=1813)

|  | N | Seroprevalence %<br>(95% CI) | p-value<sup>a</sup> | adjusted<br>p-value<sup>b</sup> | OR (95% CI)<sup>c</sup> | OR adjusted<br>p-value<sup>c</sup> |
| --- | --- | --- | --- | --- | --- | --- |
| **Period of 1+**<br>**symptoms onset** |  |  |  |  |  |  |
| No symptoms | 811 | 10.0 (8.3, 12.0) | <0.001 | <0.001 | Ref. |  |
| Feb: 1st | 150 | 14.2 (10.0, 19.7) |  |  | 1.58 (0.99, 2.51) | 0.055 |
| Feb: 2nd | 197 | 30.5 (25.1, 36.6) |  |  | 4.01 (2.83, 5.68) | <0.001 |
| March: 1st | 380 | 56.7 (52.3, 61.0) |  |  | 12.18 (9.18, 16.14) | <0.001 |
| March: 2nd | 198 | 44.2 (38.2, 50.3) |  |  | 7.03 (5.10, 9.69) | <0.001 |
| April onwards | 77 | 7.6 (3.8, 14.5) |  |  | 0.77 (0.36, 1.64) | 0.496 |
| **Period of 2+**<br>**symptoms onset** |  |  |  |  |  |  |
| At most 1 symptom | 1133 | 14.2 (12.4, 16.1) | <0.001 | <0.001 | Ref. |  |
| Feb: 1st | 103 | 14.5 (9.6, 21.2) |  |  | 1.10 (0.66, 1.83) | 0.711 |
| Feb: 2nd | 129 | 35.8 (28.9, 43.5) |  |  | 3.55 (2.46, 5.11) | <0.001 |
| March: 1st | 278 | 63.2 (58.1, 68.0) |  |  | 10.81 (8.23, 14.20) | <0.001 |
| March: 2nd | 137 | 50.3 (42.9, 57.6) |  |  | 5.93 (4.29, 8.20) | <0.001 |
| April onwards | 33 | 14.9 (7.1, 28.7) |  |  | 1.02 (0.44, 2.40) | 0.955 |

<sup>a</sup> Survey design adjusted F statistics. <sup>b</sup> Survey design adjusted F statistics, multiply adjusted for sex, age, BMI, smoking habit, main activity and municipality. <sup>c</sup> Odd ratios (OR), 95% confidence intervals (95%CI) and related adjusted p-values are obtained by linearized standard errors and logit transformation.

The most predictive symptoms were the loss of taste or smell (OR=15.05; 95%CI: 11.18, 20.26), fever (OR=5.52; 95%CI: 4.26, 7.15), difficulty in breathing (OR=5.47; 95%CI: 3.68, 8.13), pain in the limbs (OR=5.45; 95%CI: 4.35, 6.82) and weakness (OR=4.66; 95%CI: 3.66, 5.94). Symptom occurrences were visually different across age groups (Figure 2A, 2B, and 2C). In multiple logistic regression we fitted interaction terms of fever and weakness with age (Table 5). The probability of seropositivity was higher in older participants who also reported either fever or weakness (Fig. 2D). However, age was a mild predictor of infection in the absence of fever and weakness and independent of any other symptoms (OR=0.96; 95%CI: 0.93, 1.00, p=0.046; Table 5).

**Figure 2.**
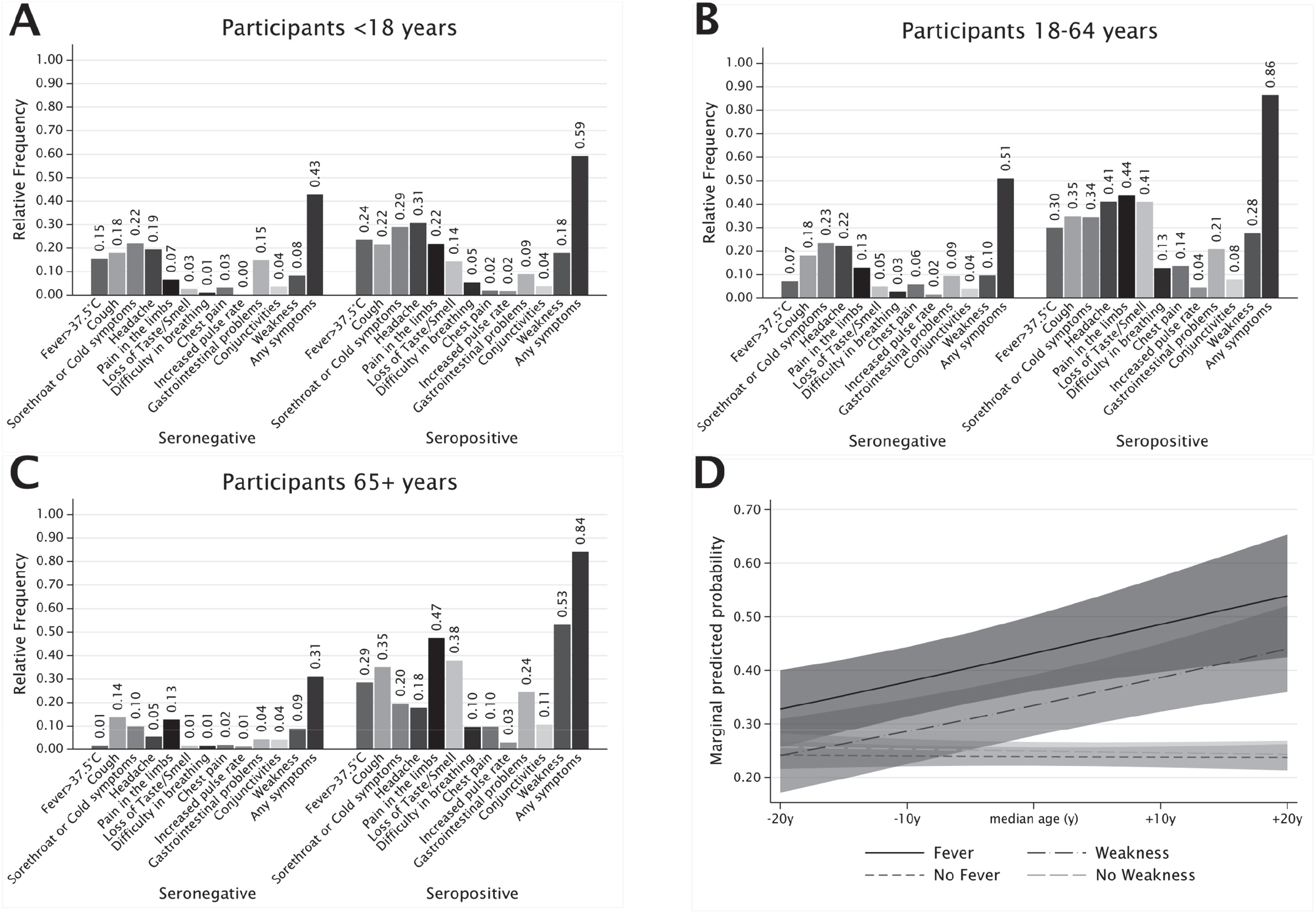
Association between reported symptoms and seroprevalence. **Panel A to C**: Symptoms frequency distribution in seronegative and seropositive participants, by age group; panel A: <18 years old; panel B: 18-64 years old; panel C: 65+ years old. **Panel D**: marginal predicted probabilities of anti SARS-CoV-2 IgG antibodies by linear effect of age as moderated by specific symptoms. Linear predictions and 95% confidence bands are displayed on the graph for participants with fever (plain line and dark grey bands), with no fever (dash line and mild grey bands), with weakness (long-dash dot line and plain grey bands) and with no weakness (long-dash line and light grey bands). For example, a participant of median age would have roughly the same marginal probability of infection of either older or younger participants, if they had no symptoms of fever and weakness, integrating across all possible predictors (e.g. Pr=0.24 if no fever present and median age, 95%CI: 0.22, 0.26). However, the estimated marginal probability of infection was 0.33 (95%CI: 0.26, 0.40) for participants 20 years younger than the median age and 0.54 (95%CI: 0.42, 0.65) for participants 20 years older than the median age, if they had fever. Corresponding probabilities for participants with weakness were 0.24 (95%CI: 0.17, 0.31) and 0.44 (95%CI: 0.36, 0.52), for participants 20 years younger and 20 years older than the median age, respectively.

**Table 5.** Results of the logistic regression model for the association between prevalence of serum antibodies to SARS-CoV-2 and symptoms while accounting for individual level relevant characteristics (n=1804).

| Predictor | Category/units | OR | 95% CI | p-value |
| --- | --- | --- | --- | --- |
| Sex | Male | Ref. |  |  |
| Sex | Female | 0.90 | (0.70, 1.15) | 0.398 |
| BMI | kg/m<sup>2</sup> | 1.04 | (1.01, 1.07) | 0.021 |
| Age | 5-yrs units | 0.96 | (0.93, 1.00) | 0.046 |
| Symptom | No symptoms | Ref. |  |  |
|  | Fever | 3.22 | (2.20, 4.71) | <0.001 |
|  | Fever X age | 1.17 | (1.07, 1.28) | 0.001 |
|  | Weakness | 1.78 | (1.23, 2.58) | 0.002 |
|  | Weakness X age | 1.19 | (1.09, 1.29) | <0.001 |
|  | Pain in the limbs | 2.76 | (2.04, 3.74) | <0.001 |
|  | Loss of taste or smell | 10.47 | (7.30, 15.01) | <0.001 |
|  | Difficulty in breathing | 1.94 | (1.07, 3.55) | 0.030 |
|  | Chest pain | 0.53 | (0.31, 0.90) | 0.018 |
| Smoking habit | Never smoker | Ref. |  |  |
|  | Past smoker | 0.96 | (0.68, 1.36) | 0.826 |
|  | Current smoker | 0.28 | (0.18, 0.44) | <0.001 |
| Main activity | Other activity | Ref. |  |  |
|  | Accommodation and catering | 1.40 | (1.02, 1.94) | 0.041 |
|  | Healthcare and social services | 1.01 | (0.55, 1.84) | 0.987 |
|  | Student or not active | 1.04 | (0.78, 1.37) | 0.812 |
| Municipality | Ortisei | Ref. |  |  |
|  | Santa Cristina | 1.34 | (0.99, 1.81) | 0.061 |
|  | Selva | 1.38 | (1.05, 1.82) | 0.020 |
| Dietary supplements | No | Ref. |  |  |
|  | Yes | 0.51 | (0.34, 0.75) | 0.001 |
| Any drug regularly | No | Ref. |  |  |
|  | Yes | 0.64 | (0.46, 0.89) | 0.008 |

Survey design adjusted odds ratios (OR) (see Methods). 95% confidence intervals (95%CI) and related adjusted p-values were obtained by linearized standard errors and logit transformation. BMI: body mass index.

Among participants with any number of symptoms (n=1002), seroprevalence peaked in those with symptoms onset in the first half of March and returned the second highest figure for onset in the following fortnight (Table 4, all p<0.001). Seroprevalence was higher also among those reporting symptoms in the second half of February, compared to other periods (p<0.001). This curvilinear trend was even more apparent when restricting the analyses to participants reporting 2+ symptoms (n=680), whereas seroprevalence peaked at 63.2% (95%CI: 58.1, 68.0) among multi-symptomatic participants with reported onset in the first half of March (Table 4, Fig. 3).

**Figure 3.**
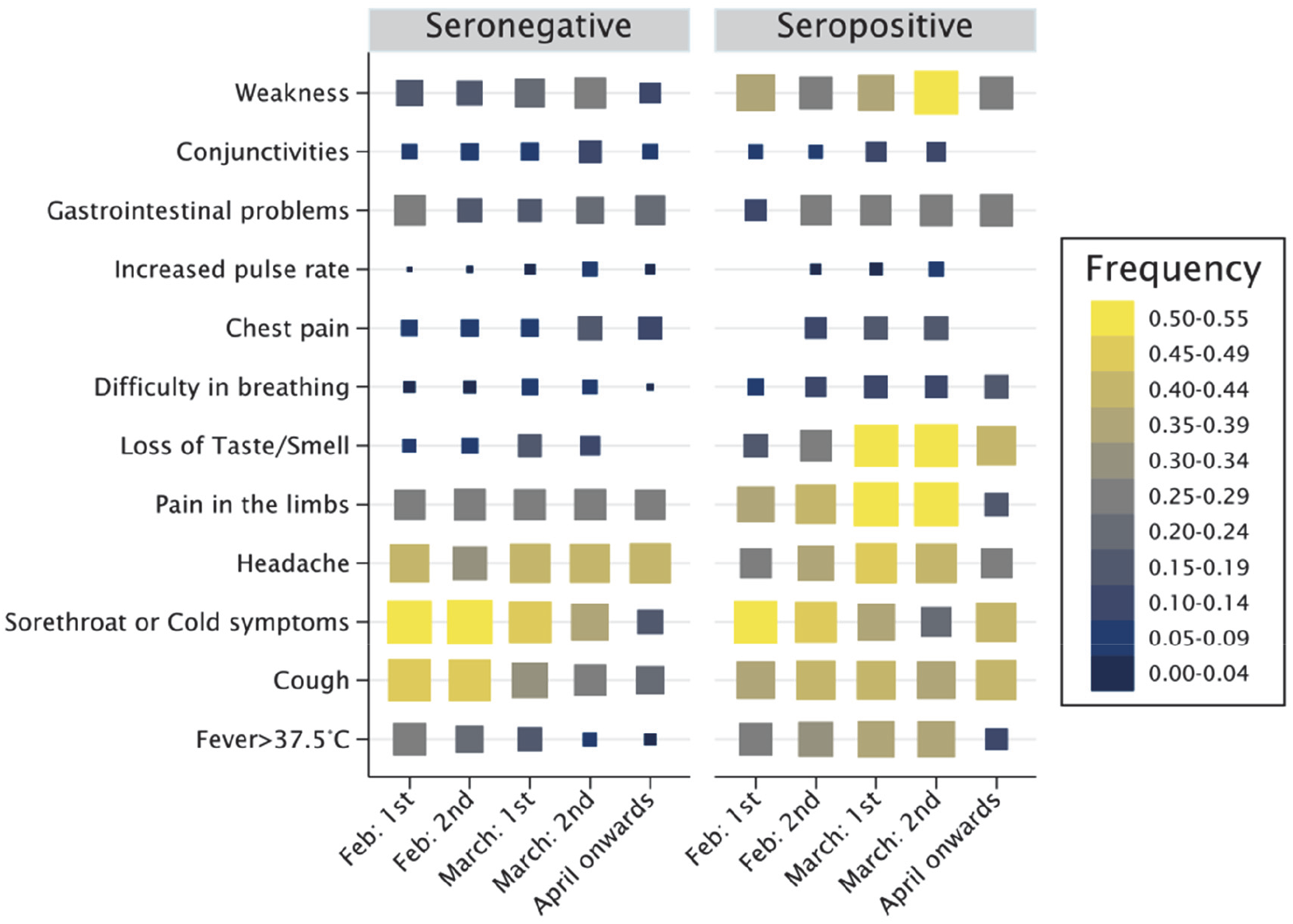
Reported symptoms by time of symptom onset in seronegative and seropositive participants. Rectangle sizes proportional to the frequency interval of symptoms across periods. The symptoms most predictive of seropositivity (loss of taste or smell; weakness; pain in the limbs; fever; and breathlessness) were apparently more prevalent among seropositive than seronegative participants at the time of peak incidence of the epidemic first wave in the valley, between late February, throughout March and part of April.

Of those participants reporting multiple symptoms (2+), 34.6% (95%CI: 31.6; 37.8) also reported having sought medical assistance (mostly by contacting the general practitioner) while 65.4% (95%CI: 62.2, 68.4) had not. Participants with any number of symptoms were more frequently testing positive (51.4%) if they had contacted the healthcare service for their symptoms than if they did not (33.9%, p<0.001), despite the minority (286 vs 716) had sought contact.

We further investigated associations with reported preexistent chronic conditions (Suppl. Tab. 4). We observed mild evidence of association with metabolic diseases (OR=0.46; 95%CI: 0.22, 0.98; p=0.043) and liver disease (OR=2.83; 95%CI: 1.00, 7.94; p=0.049) in opposite directions. As for regular therapies, directions of association were fairly consistent with those of chronic conditions (Suppl. Tab. 5). There was mild to weak evidence of negative associations of seropositivity with diabetes therapies (OR=0.38; 95%CI: 0.13, 0.92; p=0.032) and use of sedatives, antidepressants or antipsychotics (OR=0.57, 95%CI: 0.32, 1.03; p=0.064). Having had any prior health event was not associated with seropositivity (Suppl. Tab. 6). However, some reported events such as organ transplantation, chemo/radiotherapy and disautonomy were relatively infrequent to allow drawing definitive conclusions regarding these entities. Among incident health related issues, or preexisting issues worsening their clinical course after March 2020, musculoskeletal disorders (OR=2.00; 95%CI: 1.38, 2.92), respiratory or pulmonary diseases (OR=2.44; 95%CI: 1.42, 4.20), and sleep disorders (OR=1.81; 95%CI: 1.21, 2.70) were more frequent in seropositive than seronegative participants (Suppl. Tab. 6).

## DISCUSSION

The present study shows that nearly 30% of the Gardena valley general population was infected by SARS-CoV-2 during the first pandemic wave. Neutralizing antibody capacity was measurably higher than estimated seropositivity. Our in-depth analysis of SARS-CoV-2 determinants of infection as well as COVID-19-related symptoms identified specific seroprevalence risk factors and extensively characterized the post-infection symptoms.

The main strength of the study is its representativeness that guarantees generalizability of the findings to the whole population of the valley. The study was concluded in a short time span of two weeks of a nearly three-month-long strict national lockdown. The whole framework of recruitment, sample-handling, and storage procedures allowed the remarkable participation response, while preventing the possible influence of additional external factors through the period.

Our study also comes with several limitations. First, antibody testing methods have imperfect accuracy. We corrected seroprevalence analyses for the test sensitivity and specificity to overcome this limitation. Second, certain social groups such as, for instance, non-Italian residents might have been underrepresented. While we corrected all analyses for differential participation in known groups, we could not prevent participation bias of unknown sources such as, for example, COVID-19-related mortality and possible self-selection of severely ill individuals. A third limitation is the questionnaire self-administration: while this was the only way to collect essential information, response bias might affect some analyses. For instance, symptom onset estimation might not be totally accurate as the question was not specific to each possible symptom. Similarly, we cannot exclude that symptoms were due to competitive seasonal diseases such as flu or allergies.

A nearly 30% seroprevalence is a large figure compared to other studies.[4] This estimate aligns with those of nearby Italian regions.[13, 14] However, the Manaus case shows that it might still result in a non-negligible underestimation of the true infection rate.[15] Several reasons suggest that our estimate should be considered as a lower bound of the real seroprevalence. First, the survey had missed people who already died. In 2020, the raw excess all-cause mortality rate over the previous five years was between +32.8% and +72.4% in the three municipalities.[16] Second, antibodies can persist until five months or more, but we cannot exclude that IgG levels had already waned for individuals with earlier or less severe infection or less efficient immune response.[17, 18] Importantly, even in the absence of detectable IgG levels, COVID-19 patients may develop robust T-cell mediated immune response, with SARS-CoV-2 specific T cells showing a stem-like memory profile over the full disease severity spectrum.[19-21] SARS-CoV-2-specific T cells are missed by the current investigation, contributing to prevalence underestimation. Third, it is debated whether previous exposure to other coronaviruses causing the common cold could generate long-lasting SARS-CoV-2-targeting antibodies. Cross-reactive antibodies were recently identified in few adults and more frequently in children and adolescents supporting preexisting immunity.[22] Additionally, multiple studies reported cross-reactive T cell memory in 28%-50% people,[23] increasing the possibility that some preexisting immunity is already present. Fourth, adding to the extant evidence, our comparison of the Abbott SARS-CoV-2 assay against the Diasorin assay and the PRNT reflects that a relatively higher proportion of individuals may have been in contact with the SARS-CoV-2.

The imperfect agreement between the Abbott and Diasorin tests can be explained by the two assays being directed against different viral antigens with different kinetics: the anti-nucleocapsid protein IgG for Abbott and the anti-S1/S2 portions of spike protein IgG for Diasorin.[18] Furthermore, the comparison against a neutralization test identified several samples with neutralization capacity that had negative antibody test results. Part of their misclassification against the neutralization test was due to the initial definition of precautionary thresholds set by companies, where unclassifiable/dubious indices were set to negative. The recent revision of diagnostic antibody test thresholds will limit this misclassification.[24]

Standing by reported symptoms, infections have peaked during the first half of March 2020, as in neighboring regions.[3, 13, 14] Spoken language, nationality and educational level were not associated with SARS-CoV-2 seropositivity, supporting absence of social stratification in the exposure to the virus. Sex was not a major determinant of infections even though females had ∼20% lower risk than males, in line with a population study conducted in the nearby province of Trento.[14] This might reflect a higher prevention-prone behavior of females or a higher rate of neutralizing autoantibodies against type I interferon in COVID-19 severely affected males.[25] In contrast to similar studies,[13, 14] while seropositivity had no general evidence of positive association with age, it was inversely associated with age in the absence of fever and weakness in our study. This is perhaps due to a socially diverse population of positive individuals, which were younger and arguably linked to winter-sporting activities in our study. Moreover, the association of seroprevalence with municipality and the accommodation and catering services suggests that infections might have been mainly driven by unavoidable occupational circumstances.

Only a minority of symptomatic individuals sought medical advice. While experience of non-life threatening symptoms was plausible, we cannot exclude neglect or reluctance to endure quarantine by some citizens if found positive. This poses the question to the transparency and effectiveness of public authorities’ communication efforts as well as to individual behaviors.

The apparent protective effect of current smoking on SARS-CoV-2 seroprevalence is not novel.[26, 27] A collider bias effect may supersede sample representativeness.[28] COVID-19 increases mortality risk just as smoking does. Current smoking is causally related to more severe COVID-19 disease.[29] With COVID-19 and smoking intertwined to affect mortality, it is likely that the apparent protective effect of smoking on seropositivity is explained by harvesting effects on mortality, or impairment to participation by health conditions or health-prone behaviors.[30]

In conclusion, we confirm that the Gardena valley had one of the highest prevalence of SARS-CoV-2 infection in Europe. Comparisons between distinct antibody detection assays and between serum assays and serum antibodies neutralizing capacity, yet suggest an underestimation of actual seroprevalence in our study. The infection spread peculiarities in the area has probably mitigated the sex and age differences observed in other contexts. In contrast, all investigated flu-like symptoms were predictive of a positive antibody test result, with highest and cumulative evidence for the loss of taste or smell, fever, difficulty in breathing, pain in the limbs, and weakness. However, some symptoms were associated with the seroprevalence in an age-dependent mode.

Overall, findings highlight that determinants of SARS-CoV-2 infection and outcomes are context dependent, as they relate to the pattern of infection, the local population composition, and the economic dynamics. Thus prevention strategies may be tailored to the social context.

## Supporting information

STROBE checklist

## Data Availability

Any request to access the data should be forwarded to the corresponding authors.

## ACKNOWLEDGEMENTS

The authors thank Martin Matscher (Ripartizione aziendale per l’assistenza territoriale, Azienda Sanitaria dell’Alto Adige, Bolzano, Italy), Dagmar Regele (Dipartimento di Prevenzione, Servizio Igiene e Sanità pubblica, Azienda Sanitaria dell’Alto Adige, Brunico, Italy), Enrico Wegher (Direzione amministrativa, Azienda Sanitaria dell’Alto Adige, Bolzano, Italy), Marianne Siller (Direzione tecnico assistenziale, Azienda Sanitaria dell’Alto Adige, Bolzano, Italy), and Pierpaolo Bertoli (Direzione Sanitaria, Azienda Sanitaria dell’Alto Adige, Bolzano, Italy) for help in the logistics and organization, Maria Murgia (Direzione amministrativa, Azienda Sanitaria dell’Alto Adige, Bolzano, Italy) for help with privacy matters, Raffaella Binazzi (Reparto di Malattie Infettive, Azienda Sanitaria dell’Alto Adige, Bolzano, Italy) for scientific support, Christian Fuchsberger (Institute for Biomedicine, Eurac Research, Bolzano, Italy), and Roland Keim (Servizio Psicologico, Comprensorio Sanitario di Bolzano, Azienda Sanitaria dell’Alto Adige, Bolzano, Italy) for their contribution.

Preliminary results were published online: https://astat.provincia.bz.it/it/news-pubblicazioni-info.asp?news_action=4&news_article_id=641554. All authors declare no conflict.

