## Supplement for "Prevalence and determinants of serum antibodies to SARS-CoV-2 in the general population of the Gardena Valley"

### Supplementary Material

**Questionnaire** ... 2
**Swab sample analysis** ... 7
**Sample transport and storage** ... 7
**Sample selection for plaque reduction neutralization test (PRNT)** ... 7
**Supplementary Figure 1** ... 8
**Supplementary Figure 2** ... 9
**Supplementary Figure 3** ... 10
**Supplementary Table 1** ... 11
**Supplementary Table 2** ... 12
**Supplementary Table 3** ... 13
**Supplementary Table 4.** ... 14
**Supplementary Table 5** ... 15
**Supplementary Table 6** ... 16

**Questionnaire**

The questionnaire was developed ad hoc by the project team. The response options were provided in the form of checkboxes, dropdown menus, and free-text fields for numerical data such as age in years or duration of COVID-19-related symptoms in days. The questionnaire was made available in both German and Italian languages.

COVID-19 in South Tyrol: an epidemiological study

*The data will be used in pseudo-anonymized form for studies investigating COVID-19 and related pathologies. You can request the deletion of your data at any time.*

A1. Gender: ☐ Male ☐ Female

A2. Age: ______________________

A3. Mother tongue: ☐ German ☐ Italian ☐ Ladin ☐ Other language

A4. Municipality of residence: [drop-down menu] ______________________

A5. Citizenship: ☐ Italian ☐ Foreign

A6. Highest level of education:
☐ Primary school or no title
☐ Lower secondary school
☐ Vocational secondary school (2-3 years after secondary school)
☐ Upper secondary school
☐ University or a degree higher than the upper secondary school

A7. In which area are you currently employed?
☐ Agriculture and forestry, livestock
☐ Energy sector, water/wastewater, waste disposal
☐ Skilled trade, industry, installation and repair of equipment/vehicles
☐ Construction sector, public structures
☐ Commercial sector (incl. pharmacies)
☐ Transportation, magazines
☐ Food service und tourism
☐ Information and communication technology
☐ Finance and insurance
☐ Real estate sector
☐ Freelance and support services for companies, science
☐ Public administration, law enforcement agencies
☐ Education sector
☐ Health and social services (incl. facilities for small children)
☐ Housewife/Homemaker
☐ Currently seeking employment
☐ Retired
☐ Student/Pupil

A8. How many people in total, including yourself, live in your same household? _________

B1. How do you evaluate your health status in general?

| Very poor | Poor | Fair | Good | Very good |
|---|---|---|---|---|
| ☐ | ☐ | ☐ | ☐ | ☐ |

B2. Do you suffer from or have you been diagnosed with any of the following chronic diseases?
(Multiple answers possible)

☐ Lung diseases (e.g. asthma, chronic obstructive pulmonary disease)
☐ Cardiovascular diseases (e.g. coronary heart disease, atrial fibrillation, heart failure, stroke/circulation disorders in the brain)
☐ High blood pressure
☐ Kidney diseases
☐ Diseases of the immune system
☐ Tumour diseases
☐ Metabolic diseases (e.g. diabetes, excess weight)
☐ Liver diseases
☐ Mental illness, depression and/or anxiety disorder
☐ None of the above

B3. Have your health problems *worsened* or have *new ones occurred* since March 2020?
(Multiple answers possible)

☐ Musculoskeletal problems (spine, joints, muscles etc.)
☐ High blood pressure / hypertensive crisis
☐ Diabetes / blood sugar imbalances
☐ Heart troubles
☐ Respiratory or lung diseases
☐ Depressive mood / anxiety
☐ Sleeping disorders
☐ None of the above

B4. Further relevant information about you: (Multiple answers possible)

☐ Surgical procedures under general anaesthesia within the past year
☐ Radiation therapy or chemotherapy within the past year
☐ Organ transplantation during the course of your life
☐ Respiratory allergies (e.g. hay fever)
☐ Other allergies
☐ Pregnancy (currently pregnant)
☐ I am incapable of carrying out my daily activities independently
☐ None of the above

B5. Have you received the following vaccinations? (Multiple answers possible)

☐ Flu vaccination this past fall
☐ Pneumococcal vaccination in the last 12 months
☐ Other vaccinations in the last 12 months
☐ None of the above

B6. Do you take one or more of the following medications/dietary supplements regularly?
(Multiple answers possible)

☐ Aspirin (Cardioaspirina, Aspirinetta, Cardirene)
☐ Other blood-thinning medications (e.g. Sintrom, Coumadin, Eliquis, Pradaxa, Xarelto etc.)
☐ Medications for high blood pressure
☐ Medications to lower cholesterol
☐ Medications to treat diabetes
☐ Medications to treat tumour diseases
☐ Cortisone or immunosuppressive medications
☐ Anti-inflammatory drugs (e.g. Voltaren, Diclofenac, Oki, Ibuprofen)
☐ Sedatives/anti-anxiety medications, antidepressants, antipsychotics
☐ Dietary supplements (e.g. vitamins, minerals)
☐ None of the above

B7. Between 1 February 2020 and today: had you experienced, or do you have one or more of the following symptoms? (Multiple answers possible)

☐ Fever over 37,5° for at least three consecutive days
☐ Cough
☐ Sore throat or cold symptoms
☐ Headache
☐ Muscle or body aches (pain in muscles, bones and/or joints)
☐ Loss of taste or smell
☐ Difficulty breathing
☐ Chest pain or pressure
☐ Accelerated pulse
☐ Digestive problems (diarrhoea / nausea / vomiting)
☐ Conjunctivitis (pink eye / eye pain)
☐ Asthenia (weakness, generally feeling unwell)
☐ I had/have none of these symptoms

B8. For how many days did you have these symptoms? ____________________________

B9. In which time period did you experience these symptoms?

☐ First half of February
☐ Second half of February
☐ First half of March
☐ Second half of March
☐ April
☐ May
☐ June

B10. Who have you contacted to report suspicious symptoms?

☐ Family doctor
☐ Emergency Number 112
☐ Green Number 800 751 751
☐ I went directly to hospital/first aid
☐ I had symptoms but did not contact anyone
☐ I had no symptoms and therefore did not contact anyone

B11. Have you already undergone a *smear test* for the new coronavirus?
☐ Yes, with a positive result of the first smear
☐ Yes, with a negative result of the first smear
☐ Yes, but I do not know the result
☐ No

B12. Have you already undergone an *antibody test* for the new coronavirus?
☐ Yes, by means of a rapid test (blood sample from finger), with a positive result
☐ Yes, by means of a rapid test (blood sample from finger), with a negative result
☐ Yes, by means of a serological test (blood sample taken in usual manner), with a positive result
☐ Yes, by means of a serological test (blood sample taken in usual manner), with a negative result
☐ No

B13. Have you been hospitalised because of a suspected or confirmed COVID-19 infection or have you been prescribed home isolation?
☐ Yes, I was in hospital
☐ Yes, I was in home isolation
☐ Yes, I was in home isolation and in hospital
☐ Yes, in a different facility (Gossensaß or private clinic)
☐ No

B14. Do you smoke or have you smoked in the past?
☐ I have never smoked (or *less* than 100 cigarettes in my life)
☐ Not currently, but I used to smoke
☐ Yes, I smoke occasionally (not daily)
☐ Yes, I smoke less than 10 cigarettes per day
☐ Yes, I smoke 10 to 20 cigarettes per day
☐ Yes, I smoke more than 20 cigarettes per day

B15. Your height (in cm): ________________________________________
B16. Your weight (in kg): ________________________________________

C1. Between 9 March (Government Emergency Decree) and 3 May 2020: how did you practice your profession?
☐ I continued to work at my usual place of employment
☐ from home/Smart work
☐ I had to interrupt my work activity due to the emergency
☐ I do not work, and I did not work in February either
☐ I was transferred to the wage compensation fund

C2. Between 9 March 2020 and 3 May 2020: how often did you use public transportation during this time period?
____________ days per week, on average

C3. Between 9 March,2020 und 3 May 2020: with which people were you in close contact? (except household)
☐ with no one
☐ with close family members
☐ with other relatives
☐ with work colleagues and/or superiors
☐ with dependent persons (e.g. people in need of care)
☐ with friends
☐ others: ___________________________________________________

C4. How many people were you in close contact with during this period? (except household) __________

C5. Were you in close contact (less than 2 m distance, enclosed rooms such as home, workplace, transportation) with *confirmed* COVID-19-infected persons?

☐ Yes ☐ No ☐ Don't know

C6. Were you in close contact (less than 2 m distance, enclosed rooms such as home, workplace, transportation) with *suspected* COVID-19-cases?

☐ Yes ☐ No ☐ Don't know

#### Swab sample analysis

Swab samples were analyzed at the ÖNORM-accredited (EN ISO 15189:2013) diagnostic laboratory of the Institute of Virology of the Innsbruck Medical University (IVIMU, Austria), after being kept in virus transport medium (2 mL, Hanks Balanced Salt Solution with 2% fetal bovine serum, 100 µg/ml Gentamicin, 0.5 µg/ml Fungizone) until testing. For RNA extraction, 50 µl of the inoculated virus transport medium was incubated with an in-house direct lysis buffer (10 mM TRIS-HCl pH 7.4, 25 nM NaCl, 0.5% IGEPAL, 10 Units RiboLock RNase Inhibitor in DEPC-treated Water) for 20 minutes at room temperature. Ten µl RNA eluates were used in a 20 µl RealStar® SARS-CoV-2 RT-PCR (Altona-Diagnostics GmbH, Hamburg, Germany) master mix on the CFX96TM RT-PCR Detection System (Bio-Rad Laboratories, Inc.). To confirm positive samples, RNA was extracted again from the original material using the EasyMag® NucliSENS® System (Biomérieux Deutschland, Nürtingen, Germany) and used for RT-PCR as described above.

#### Sample transport and storage

Biological samples (serum of 6+ year old participants; serum and whole blood of 18+ year old participants) were transferred from the Bressanone/Brixen Hospital to the Eurac Research biobank at the Bolzano/Bozen Hospital at 4°C controlled temperature. The serological analysis leftover was split into up-to-4 250 µl aliquots (Thermo Scientific™ Nunc™ Biobanking Tubes, Ref. 374086, Thermo Fisher Scientific) using a Hamilton Robotics STARlet liquid handler. Samples were frozen by direct immersion in liquid nitrogen and stored at -80°C together with an additional 3 ml whole blood/EDTA tube (*VACUETTE*®, Ref. 454411, Greiner).

#### Sample selection for plaque reduction neutralization test (PRNT)

From the 2129 samples with available serological test results, we selected 299 samples to submit to PRNT (sample size defined by budgetary and logistical constraints). To maximize the utility of the 299 samples from the 2129 individuals, we subset the 1461 ones with maximal storage availability (4 cryo-conserved aliquots) and valid response to both questions B11 "*Have you already undergone a smear test for the new coronavirus?*" and B13 "*Have you been hospitalised because of a suspected or confirmed COVID-19 infection or have you been prescribed home isolation?*" (see Questionnaire above, page 5). To select the most informative subset of 299 samples out of the available 1461 ones, we split the 1461 observations into 280 quantiles using the *ntiles()* function implemented in the R package 'schoRsch' version 1.8[1] guaranteeing at least 3 available samples per bin. Within each bin, we aimed at selecting the combination of samples maximizing the information content of the corresponding epidemiological database. We implemented an iterative algorithm that, each time, sampled 299 samples (1 or 2 per bin) and calculated an entropy index on the corresponding dataset from the questionnaires' responses. Each time, if the entropy value estimated from the newly extracted 299 samples was larger than the previously estimated one, then the new set was retained, otherwise, it was discarded. Entropy was estimated using a shrinkage method based on a Dirichlet distribution as implemented in the *entropy()* function in the R package "infotheo" version 1.2.0 (https://cran.r-project.org/package=infotheo). We performed 50,000 resamplings. Sensitivity analyses using alternative methods gave identical results, namely: the same 299 selected samples. **Suppl. Fig. 1A** shows that the maximal entropy was achieved after about 5000 iterations, making it unlikely that a better configuration could arise beyond the 50,000$^{th}$ iteration. Entropy distribution is given in **Suppl. Fig. 1B**. In terms of S/C (AAI) values, the 299 selected samples were evenly distributed over the whole available range of the 1461 starting samples (**Suppl. Fig. 1C**). Analyses were performed using the software R version 3.6.3 (www.r-project.org).

**Supplementary Figure 1**. Sample selection for neutralization capacity test. **Panel A**: estimation of the entropy value for each of 50,000 iterations: the early stabilization suggests that it is unlikely to identify a more informative sample with more iterations. **Panel B**: entropy distribution in the 299 selected samples. **Panel C**: Levels of the 299 selected samples in S/C (AAI) units (white dots) in the context of the 1461 starting samples (black dots), sorted by increasing value: selected samples cover well the original distribution.

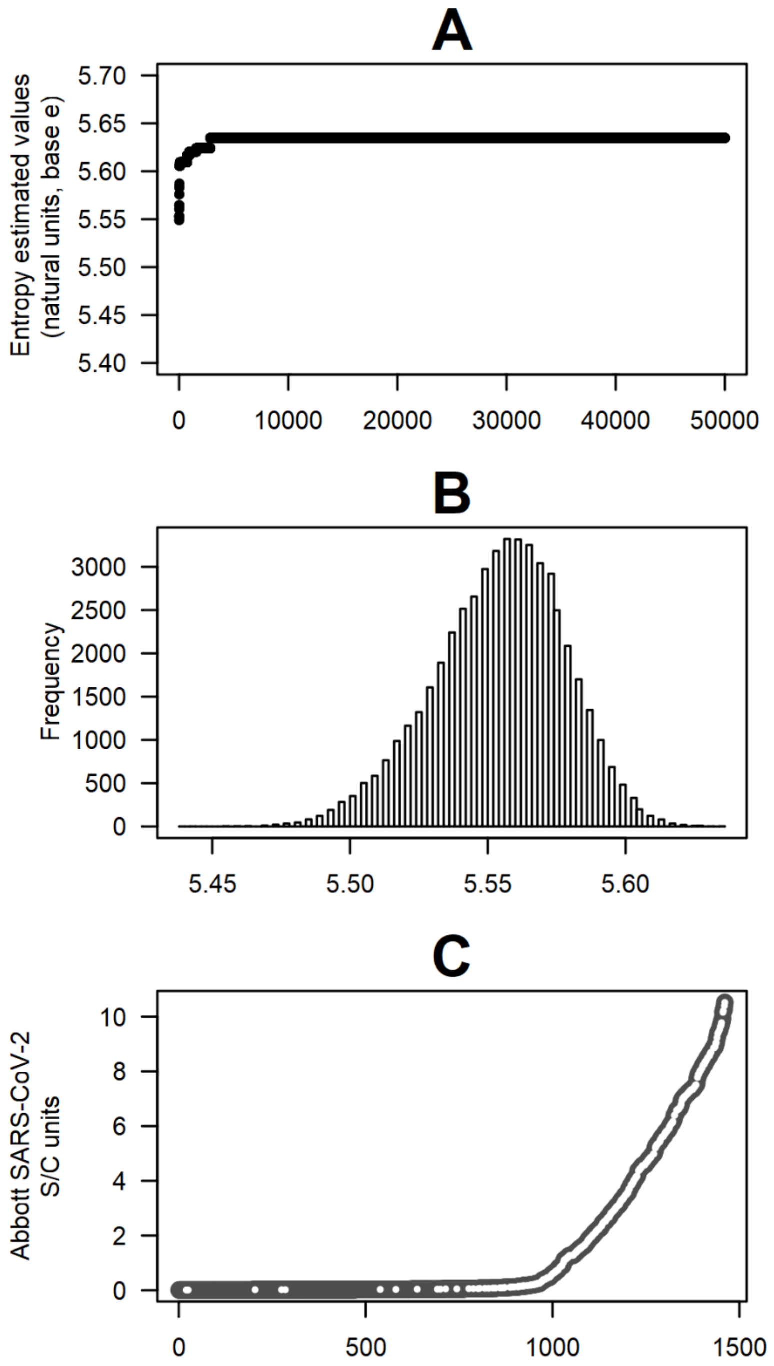

**Supplementary Figure 2**. Concordance between S/C (AAI) levels from the Abbott test on fresh serum and thawed serum after freezing, evaluated using Lin's concordance correlation coefficient (CCC)[2] and Bland & Altman (B&A)'s plot.[3] **Panel A**: The linear regression fitted line overlapping the bisecting line and the very high Lin's CCC estimated coefficient indicate almost perfect concordance. **Panel B**: B&A's plot showing mean S/C (AAI) units across the two measurements (x-axis) versus the difference of values on fresh versus thawed serum: white dots indicate samples for which a slightly lipemic, lipemic, slightly haemolitic, or haemolotic status was observed (when removing these samples, Lin's CCC = 0.9986, 95%CI: 0.9977, 0.9992); blue dots identify the few samples that were stored for >1 day (up to 3 days) before processing and freezing (when removing these samples, Lin's CCC was 0.9985, 95%CI: 0.9982, 0.9992). **Panel C**: B&A's plots by plate: while plate effect explained ~13% of the differences between S/C values measured by Abbott test on fresh versus thawed serum, within-plate concordance was very high (Lin's CCC >0.99 for every plate).

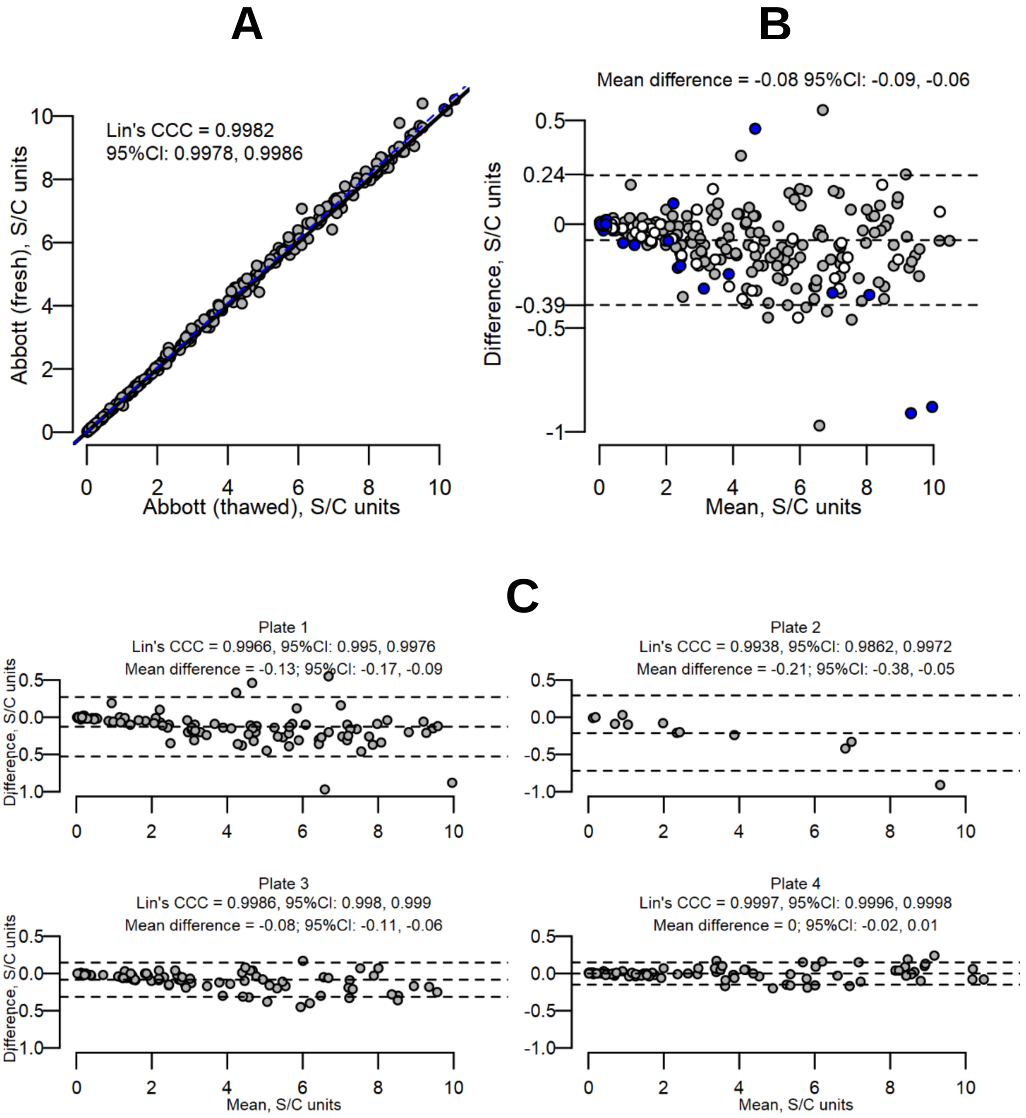

**Supplementary Figure 3**. ROC curve sub-group analyses. The cross symbol corresponds to the classifier performance at the 1.40 canonical S/C (AAI) threshold (Abbott assay). The optimal classifier based on the Youden's index (*J*) may vary and is displayed with a circle in each curve. The thresholds obtained from 10-fold cross validation and repeated random sub-sampling validation differ from the thresholds obtained from the respective sub-groups shown in these plots, limiting their reliability and applicability. **Panel A**: Sex groups. Black and dark gray lines represent the ROC curves obtained for females (n=140) and males (n=159), respectively. Of note, restriction to males yields no false positives for the canonical 1.40 threshold. **Panel B**: Age groups. Black and dark gray lines represent participants ≥ 48 years of age (n=153) and <48 years of age (n=146), respectively. We observe that the classifier has relatively higher specificity when applied to participants at least 48 years of age. See Supplementary Table 3 for more details.

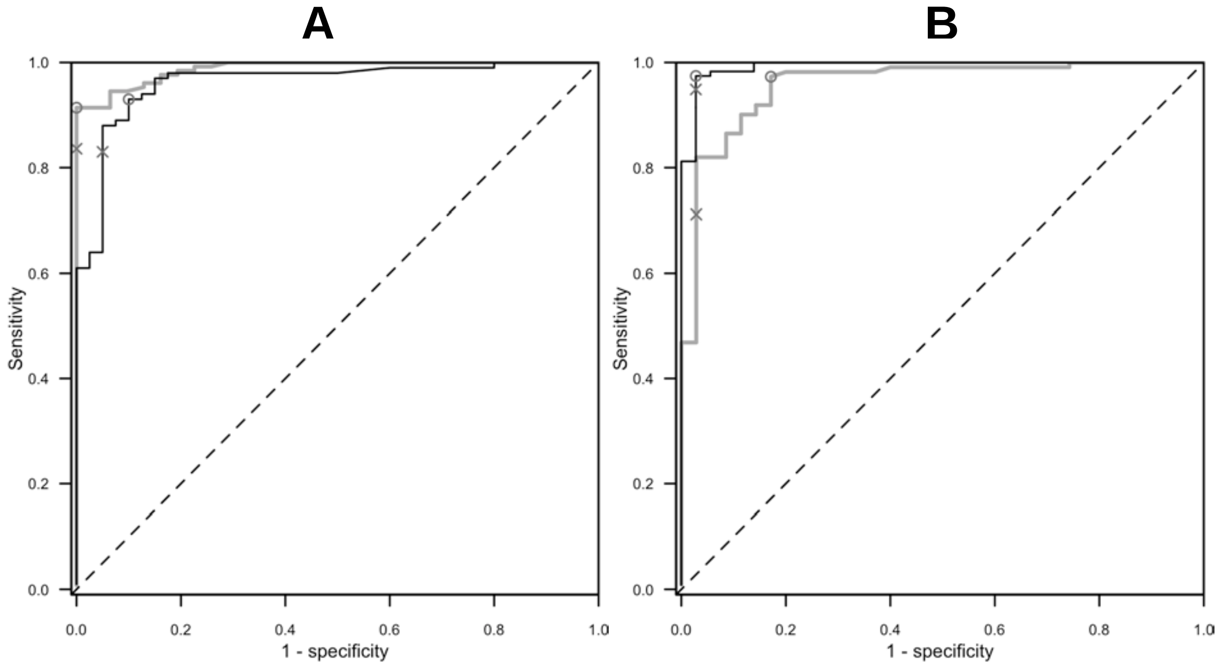

**Supplementary Table 1**. Prevalence of serum antibodies to SARS-CoV-2 by participant's questionnaire completion status and overall seroprevalence.

| | Design-based seroprevalence | | | |
|---|---|---|---|---|
| **Questionnaire complete** | **No (%)** (n=1555) | **Yes (%)** (n=551) | ***P* value**[a] | **Adjusted seroprevalence (95%CI)**[b] |
| No (n=293) | 72.1% | 27.9% | 0.447 | 28.6% (24.1%, 33.6%) |
| Yes (n=1813) | 74.0% | 26.0% | | 26.6% (24.8%, 28.5%) |
| **Overall (n=2106)** | **73.7%** | **26.3%** | | 26.9% (25.2%, 28.6%) |

Seroprevalence estimate based on Abbott SARS-CoV-2 IgG antibody test, cutoff 1.4. Participants were aged 6+, pregnant women were excluded, and women with valid serological test results but who didn’t respond to the questionnaire were assumed not to be pregnant. Tabulated proportions account for the survey design settings. ‘n’ counts are actual marginal observations.

[a] Pearson design based F(1, 2083) test for difference between proportions.

[b] Estimates account for the survey design settings (see Methods) and are adjusted to the serological test accuracy (sensitivity 0.9677; specificity 0.9963). 95%CI are obtained by the logit transformation.

**Supplementary Table 2**. Distribution of the 299 samples submitted to plaque reduction neutralization test (PRNT).

| **Quantitative variables** | | **Min** | **Q1, Median, Q3** | **Max** | **Mean(SD)** |
|---|---|---|---|---|---|
| Age, years | | 19.0 | 32.0, 48.0, 92.0 | 92.0 | 47.3 (18.1) |
| Results of Abbott IgG assay on fresh serum, S/C units | | 0.01 | 0.60, 5.75, 10.52 | 10.52 | 3.44 (2.98) |
| Results of Abbott IgG assay on thawed serum, S/C units | | 0.02 | 0.60, 2.67, 5.63 | 10.44 | 3.36 (2.93) |
| Results of Diasorin assay on thawed serum, AUs units | | 3.80 | 5.15, 26.80, 57.80 | 400.00 | 49.20 (67.96) |
| **Categorical variables** | **Level** | **n (%)** | | | |
| Sex | Female | 140 (46.8) | | | |
| | Male | 159 (53.2) | | | |
| Plate number | 1 | 96 (32.1) | | | |
| | 2 | 12 (4.0) | | | |
| | 3 | 95 (31.8) | | | |
| | 4 | 96 (32.1) | | | |
| Days between sample collection and processing | 1 | 280 (93.6) | | | |
| | 3 | 19 (6.4) | | | |
| Previous swab test | No | 269 (90.0) | | | |
| | Yes, Negative | 19 (6.4) | | | |
| | Yes, not known | 2 (0.7) | | | |
| | Yes, Positive | 9 (3.0) | | | |
| Previous serological test (needle or pinprick) | No | 220 (73.6) | | | |
| | Yes, Negative | 23 (7.7) | | | |
| | Yes, Positive | 56 (18.7) | | | |
| Abbott test ≥1.4, fresh serum | Negative | 105 (35.1) | | | |
| | Positive | 194 (64.9) | | | |
| Abbott test ≥1.4, thawed serum | Negative | 107 (35.8) | | | |
| | Positive | 192 (64.2) | | | |
| Diasorin test | Negative | 102 (34.1) | | | |
| | Inconclusive | 7 (2.3) | | | |
| | Positive | 190 (63.5) | | | |
| NT titer 50% | Negative | 71 (23.7) | | | |
| | Positive | 228 (76.3) | | | |
| NT titer 90% | Negative | 134 (44.8) | | | |
| | Positive | 165 (55.2) | | | |

*Abbreviations*: Q1, 1st quartile; Q3, 3rd quartile; NT titer, neutralizing titer.

**Supplementary Table 3**. Discrimination accuracy of the Abbott antibody assay versus the serum neutralization capacity, by ROC curve analysis in the whole sample and in sub-groups.

| Variable | Group | N[a] | AAI[b] | Sensitivity | Specificity |
|---|---|---|---|---|---|
| (none) | Whole sample | 299 | 1.40 | 83.3% | 92.7% |
| | | | 1.16 | 89.0% | 97.2% |
| Sex | Male | 159 | 1.40 | 83.6% | 100.0% |
| | | | 1.16 | 91.4% | 100.0% |
| | Female | 140 | 1.40 | 83.0% | 95.0% |
| | | | 0.54 | 93.0% | 90.0% |
| Age | <48 years | 146 | 1.40 | 71.2% | 97.1% |
| | | | 0.36 | 97.3% | 82.9% |
| | ≥48 years | 153 | 1.40 | 94.9% | 97.2% |
| | | | 1.00 | 97.4% | 97.2% |

[a] Number of participants in each group.
[b] S/C (AAI) threshold used to derive sensitivity and specificity. Corresponds to either the Abbott's suggested cutoff AAI = 1.40 (marked by a cross in the ROC curves) or to the optimal cutoff derived by ROC analysis (marked by a circle).

**Supplementary Table 4.** Prevalence of serum antibodies to SARS-CoV-2 by participant's reported chronic disease condition[a].

| Chronic disease | Category | N | Seroprevalence% (95% CI) | F statistic p-value[b] | OR (95% CI)[c] | p-value[c] |
|---|---|---|---|---|---|---|
| Pulmonary Disease | No | 1771 | 26.1 (24.3; 27.9) | 0.478 | Ref. | |
| | Yes | 42 | 21.8 (12.8; 34.8) | | 0.76 (0.39; 1.48) | 0.418 |
| Cardiovascular disease | No | 1740 | 26.0 (24.2; 27.8) | 0.956 | Ref. | |
| | Yes | 73 | 26.2 (18.3; 36.0) | | 0.92 (0.57; 1.50) | 0.744 |
| Hypertension | No | 1647 | 26.1 (24.3; 28.0) | 0.682 | Ref. | |
| | Yes | 166 | 24.8 (19.6; 31.0) | | 0.82 (0.58; 1.17) | 0.270 |
| Kidney Disease | No | 1799 | 25.9 (24.2; 27.8) | 0.415 | Ref. | |
| | Yes | 14 | 34.3 (16.6; 57.8) | | 1.48 (0.54; 4.05) | 0.449 |
| Autoimmune Disease | No | 1792 | 25.9 (24.1; 27.7) | 0.173 | Ref. | |
| | Yes | 21 | 37.3 (21.6; 56.1) | | 1.97 (0.88; 4.41) | 0.098 |
| Tumor | No | 1758 | 26.1 (24.3; 28.0) | 0.519 | Ref. | |
| | Yes | 55 | 22.7 (14.5; 33.7) | | 0.87 (0.49; 1.53) | 0.622 |
| Metabolic Disease | No | 1767 | 26.3 (24.5; 28.1) | 0.053 | Ref. | |
| | Yes | 46 | 15.2 (8.1; 26.6) | | 0.46 (0.22; 0.98) | 0.043 |
| Liver Disease | No | 1803 | 25.9 (24.1; 27.7) | 0.034 | Ref. | |
| | Yes | 10 | 51.6 (26.4; 75.9) | | 2.83 (1.00; 7.94) | 0.049 |
| Psychosis / Affective Disorder | No | 1773 | 26.2 (24.4; 28.1) | 0.109 | Ref. | |
| | Yes | 40 | 16.6 (8.9; 28.8) | | 0.60 (0.29; 1.25) | 0.169 |
| Any chronic condition | No | 1443 | 26.3 (24.3; 28.3) | 0.546 | Ref. | |
| | Yes | 370 | 24.9 (21.3; 29.0) | | 0.87 (0.67; 1.13) | 0.308 |

[a] Prevalence estimate based on Abbott SARS-CoV-2 IgG antibody test, cutoff 1.4.
[b] Survey design adjusted F statistics.
[c] Odd ratios (OR) adjusted for gender, age, body mass index, smoking habit, activity-group, and municipality; 95% confidence intervals (95%CI) and p-values obtained by linearized standard errors and logit transformation.

**Supplementary Table 5.** Prevalence of serum antibodies to SARS-CoV-2 by participant's reported regular therapy[a].

| **Therapy** | **Category** | **N** | **seroprevalence% (95% CI)[a]** | **F statistic p-value[b]** | **OR (95% CI)[c]** | **p-value[c]** |
|---|---|---|---|---|---|---|
| Aspirin (incl. cardioaspirin) | No | 1732 | 26.0 (24.2; 27.9) | 0.888 | Ref. | - |
| | Yes | 81 | 25.4 (18.0; 34.5) | | 1.02 (0.63; 1.64) | 0.936 |
| Anticoagulants | No | 1745 | 26.0 (24.2; 27.8) | 0.923 | Ref. | - |
| | Yes | 68 | 26.4 (18.3; 36.5) | | 0.97 (0.58; 1.63) | 0.903 |
| For Hypertension | No | 1611 | 25.9 (24.1; 27.8) | 0.747 | Ref. | - |
| | Yes | 202 | 26.8 (21.8; 32.5) | | 0.96 (0.69; 1.34) | 0.825 |
| For cholesterol | No | 1755 | 25.9 (24.2; 27.8) | 0.750 | Ref. | - |
| | Yes | 58 | 27.6 (18.7; 38.6) | | 0.96 (0.57; 1.62) | 0.883 |
| For Diabetes | No | 1775 | 26.3 (24.5; 28.1) | 0.037 | Ref. | - |
| | Yes | 38 | 13.1 (6.2; 25.6) | | 0.38 (0.16; 0.92) | 0.032 |
| Anticancer | No | 1801 | 26.1 (24.4; 27.9) | 0.110 | Ref. | - |
| | Yes | 12 | 8.3 (1.5; 35.2) | | 0.32 (0.06; 1.86) | 0.206 |
| Cortisone / immunosuppr. therapy | No | 1786 | 26.0 (24.3; 27.8) | 0.910 | Ref. | - |
| | Yes | 27 | 25.2 (13.6; 41.8) | | 0.98 (0.47; 2.04) | 0.947 |
| Antiinflammatory (NSAIDs) | No | 1726 | 26.1 (24.3; 27.9) | 0.758 | Ref. | - |
| | Yes | 87 | 24.8 (17.7; 33.5) | | 0.96 (0.63; 1.48) | 0.867 |
| Sedatives, antidepressants or anti-psychotics | No | 1752 | 26.3 (24.5; 28.2) | 0.060 | Ref. | - |
| | Yes | 61 | 17.1 (10.5; 26.7) | | 0.57 (0.32; 1.03) | 0.064 |
| Dietary supplements | No | 1613 | 26.2 (24.3; 28.1) | 0.533 | Ref. | - |
| | Yes | 200 | 24.4 (19.6; 30.0) | | 0.90 (0.66; 1.23) | 0.524 |
| Any drug regularly (excl. dietary) | No | 1370 | 26.4 (24.4; 28.6) | 0.385 | Ref. | - |
| | Yes | 443 | 24.6 (21.3; 28.3) | | 0.83 (0.64; 1.08) | 0.170 |

[a] Prevalence estimate based on Abbott SARS-CoV-2 IgG antibody test, cutoff 1.4.
[b] Survey design adjusted F statistics.
[c] Odd ratios (OR) adjusted for sex, age, body mass index, smoking habit, activity-group and municipality; 95% confidence intervals (95%CI) and p-values obtained by linearized standard errors and logit transform.

**Supplementary Table 6.** Prevalence of serum antibodies to SARS-CoV-2 by participant's questionnaire responses to prior health events, vaccinations, and post 9$^{th}$ March 2020 health conditions.

| **Health issues** | **Type** | **category** | **N** | **seroprevalence% (95% CI)** | **F statistic p-value[a]** | **OR (95% CI)[b]** | **p-value[b]** |
|---|---|---|---|---|---|---|---|
| Health events | Surgical Intervention | No | 1700 | 26.1 (24.3; 28.0) | 0.259 | Ref. | |
| | | Yes | 113 | 24.7 (18.4; 32.3) | | 0.80 (0.55; 1.18) | 0.259 |
| | Chemo/Radio-therapy | No | 1803 | 26.1 (24.3; 27.9) | 0.366 | Ref. | |
| | | Yes | 10 | 10.5 (1.9; 41.6) | | 0.42 (0.07; 2.74) | 0.366 |
| | Organ Transplant | No | 1809 | 26.0 (24.3; 27.8) | 0.914 | Ref. | |
| | | Yes | 4 | 22.8 (4.1; 67.1) | | 0.91 (0.15; 5.46) | 0.914 |
| | Respiratory allergy | No | 1601 | 26.3 (24.4; 28.2) | 0.385 | Ref. | |
| | | Yes | 212 | 23.9 (19.2; 29.3) | | 0.87 (0.64; 1.19) | 0.385 |
| | Other allergy | No | 1743 | 26.4 (24.6; 28.2) | 0.112 | Ref. | |
| | | Yes | 70 | 17.5 (10.7; 27.3) | | 0.61 (0.34; 1.12) | 0.112 |
| | Disautonomy | No | 1796 | 26.1 (24.3; 27.9) | 0.549 | Ref. | |
| | | Yes | 17 | 17.7 (6.7; 39.1) | | 0.71 (0.23; 2.20) | 0.549 |
| | Major Health Event | No | 1677 | 26.1 (24.3; 28.0) | 0.293 | Ref. | |
| | | Yes | 136 | 24.2 (18.5; 31.0) | | 0.83 (0.58; 1.18) | 0.293 |
| | Any allergy | No | 1551 | 26.5 (24.6; 28.5) | 0.247 | Ref. | |
| | | Yes | 262 | 23.2 (19.0; 28.1) | | 0.84 (0.63; 1.12) | 0.247 |
| Type of vaccine | Seasonal Flu | No | 1629 | 25.8 (24.0; 27.7) | 0.528 | Ref. | |
| | | Yes | 184 | 27.6 (22.4; 33.6) | | 1.06 (0.76; 1.49) | 0.727 |
| | Pneumococcus | No | 1801 | 26.0 (24.3; 27.8) | 0.855 | Ref. | |
| | | Yes | 12 | 24.1 (9.2; 49.8) | | 1.15 (0.36; 3.71) | 0.812 |
| | Other vaccination | No | 1723 | 26.3 (24.5; 28.1) | 0.227 | Ref. | |
| | | Yes | 90 | 21.0 (14.3; 29.8) | | 0.76 (0.46; 1.25) | 0.281 |
| | Any vaccination | No | 1537 | 26.2 (24.3; 28.2) | 0.640 | Ref. | |
| | | Yes | 276 | 25.0 (20.8; 29.7) | | 0.92 (0.70; 1.22) | 0.560 |
| Post 9th March health condition | Incident / worse muskuloskeletal disease | No | 1716 | 25.2 (23.4; 27.1) | <0.001 | Ref. | |
| | | Yes | 97 | 40.6 (32.4; 49.4) | | 2.00 (1.38; 2.92) | <0.001 |
| | Incident / worse hypertension / elevated blood pressure | No | 1796 | 25.9 (24.2; 27.7) | 0.340 | Ref. | |
| | | Yes | 17 | 34.7 (18.3; 55.7) | | 1.61 (0.66; 3.90) | 0.292 |
| | Incident / worse diabetes / hyperglycemia | No | 1810 | 26.0 (24.2; 27.8) | 0.604 | Ref. | |
| | | Yes | 3 | 37.9 (6.9; 83.5) | | 1.70 (0.22; 13.27) | 0.612 |
| | Incident / worse cardiac disease | No | 1798 | 26.1 (24.3; 27.9) | 0.223 | Ref. | |
| | | Yes | 15 | 13.8 (4.2; 37.1) | | 0.43 (0.12; 1.60) | 0.209 |
| | Incident / worse respiratory / pulmonary disease | No | 1771 | 25.5 (23.7; 27.3) | <0.001 | Ref. | |
| | | Yes | 42 | 46.7 (33.9; 60.0) | | 2.44 (1.42; 4.20) | 0.001 |
| | Incident / worse depression / phobia | No | 1784 | 25.9 (24.1; 27.7) | 0.360 | Ref. | |
| | | Yes | 29 | 32.5 (19.6; 48.6) | | 1.67 (0.80; 3.52) | 0.174 |
| | Incident / worse sleep disorders | No | 1721 | 25.4 (23.6; 27.2) | 0.004 | Ref. | |
| | | Yes | 92 | 37.6 (29.2; 46.7) | | 1.81 (1.21; 2.70) | 0.004 |

[a] Survey design adjusted F statistics.
[b] Odd ratios (OR) adjusted for gender, age, body mass index, smoking habit, activity-group and municipality. 95% confidence intervals (95%CI) and p-values obtained by linearized standard errors and logit transformation.

[c] Major health event includes any of: surgical intervention, chemo/radio therapy, organ transplant or disautonomy.
