## Supplementary material for "Prevalence and determinants of serum antibodies to SARS-CoV-2 in the general population of the Gardena Valley": STROBE checklist

STROBE Statement—Checklist of items that should be included in reports of ***cross-sectional studies***

| | Item No | Recommendation |
|---|---|---|
| **Title and abstract** | 1 | ☒(*a*) Indicate the study's design with a commonly used term in the title or the abstract *Abstract identifies the study as a cross-sectional study* |
| | | ☒(*b*) Provide in the abstract an informative and balanced summary of what was done and what was found *Abstract characterizes the cross-sectional study, provides information on antibody bioassays used to determine seroprevalence, and discusses statistical methodology and findings.* |
| **Introduction** | | |
| Background/rationale | 2 | ☒Explain the scientific background and rationale for the investigation being reported *SARS-CoV-2 seroprevalence study in a geographically restricted region with high infection incidence.* |
| Objectives | 3 | ☒State specific objectives, including any prespecified hypotheses *Characterization of SARS-CoV-2 seroprevalence in the valley population. Observational study without construction of prior hypotheses.* |
| **Methods** | | |
| Study design | 4 | ☒Present key elements of study design early in the paper *Biology-related methods are addressed first, followed by computational methods. Items 4, 5, and 6 are addressed in the first paragraph of the Methods section.* |
| Setting | 5 | ☒Describe the setting, locations, and relevant dates, including periods of recruitment, exposure, follow-up, and data collection *Population size and number of invited participants given immediately.* |
| Participants | 6 | ☒(*a*) Give the eligibility criteria, and the sources and methods of selection of participants *Sample selection is described; eligibility criteria: place of residence; age ≥ 6; non-pregnant women* |
| Variables | 7 | ☒Clearly define all outcomes, exposures, predictors, potential confounders, and effect modifiers. Give diagnostic criteria, if applicable *Derivation of descriptive variables, definition of strata and use of regression tests are addressed in a section on statistical analyses. Measurement of SARS-CoV-2 exposure is detailed in the biology section.* |
| Data sources/ measurement | 8* | ☒For each variable of interest, give sources of data and details of methods of assessment (measurement). Describe comparability of assessment methods if there is more than one group *Variables are obtained from a questionnaire, which is available as a supplement. SARS-CoV-2 exposure is measured by using nasopharyngeal swab and serum antibody tests.* |
| Bias | 9 | ☒Describe any efforts to address potential sources of bias *Various types of possible bias were accounted for in the Methods, both at the study design phase and in the post collection data analyses (see Statistical analyses section). The Discussion section discusses additional potential sources of bias that could not be accounted for.* |
| Study size | 10 | ☒Explain how the study size was arrived at *Sample size determination is described in the first paragraph of the Methods section.* |
| Quantitative | 11 | ☒Explain how quantitative variables were handled in the analyses. If applicable, |

| | | |
|---|---|---|
| variables | | describe which groupings were chosen and why<br>*When applicable, manufacturers thresholds were chosen for deriving groupings and any deviations are clearly marked and discussed. Age groups include non-adults and elderly as separate strata.* |
| Statistical methods | 12 | ☒(*a*) Describe all statistical methods, including those used to control for confounding<br>*Descriptive statistics, derivation of confidence intervals and regression tests are detailed in the Methods (Statistical analyses section) and table footers.* |
| | | ☒(*b*) Describe any methods used to examine subgroups and interactions<br>*Predictors in logistic regression models make use of interactions, and a separate graph is dedicated to their interpretation. ROC analyses on serum antibodies split subroups by age and sex in separate graphs* |
| | | ☒(*c*) Explain how missing data were addressed<br>*Missing data were addressed by using non-response corrected sampling weights in all analyses.* |
| | | ☒(*d*) If applicable, describe analytical methods taking account of sampling strategy<br>*Logistic regression models were survey set based on sample stratification and weight adjusted accounting also for post-stratification variables.* |
| | | ☒(*e*) Describe any sensitivity analyses<br>*We controlled if there was any evidence of a different estimated seroprevalence between completers and non-completers of the questionnaire interview.* |
| **Results** | | |
| Participants | 13* | ☒(a) Report numbers of individuals at each stage of study—eg numbers potentially eligible, examined for eligibility, confirmed eligible, included in the study, completing follow-up, and analysed<br>*Numbers are clearly stated.* |
| | | ☒(b) Give reasons for non-participation at each stage<br>*Reasons for non-participation are listed.* |
| | | ☒(c) Consider use of a flow diagram<br>*Given the workflow is straightforward, we explained it in the text.* |
| Descriptive data | 14* | ☒(a) Give characteristics of study participants (eg demographic, clinical, social) and information on exposures and potential confounders<br>*The first part of the Results section and Table 1 provide all descriptive data.* |
| | | ☒(b) Indicate number of participants with missing data for each variable of interest<br>*Missingness for the main outcomes is reported at the beginning of the results. Missingness for any variable of interest is indicated in the results tables or was accounted for in the statistical models* |
| Outcome data | 15* | ☒Report numbers of outcome events or summary measures<br>*Table 2 provides these data.* |
| Main results | 16 | ☒(*a*) Give unadjusted estimates and, if applicable, confounder-adjusted estimates and their precision (eg, 95% confidence interval). Make clear which confounders were adjusted for and why they were included<br>*To be found in Tables 3 and 4.* |
| | | ☒(*b*) Report category boundaries when continuous variables were categorized<br>*When continuous variables were categorized, boundaries are given* |
| | | ☐(*c*) If relevant, consider translating estimates of relative risk into absolute risk for a meaningful time period<br>*Not applicable.* |

| | | |
|---|---|---|
| Other analyses | 17 | ☒Report other analyses done—eg analyses of subgroups and interactions, and sensitivity analyses<br>*Regression models, concordance/agreement analyses, and receiver operating characteristic curve analysis are described and presented in the Methods, Supplementary Materials, and Results.* |
| **Discussion** | | |
| Key results | 18 | ☒Summarise key results with reference to study objectives<br>*All key results are summarized in the first paragraph of the Discussion and discussed in more detail in the following* |
| Limitations | 19 | ☒Discuss limitations of the study, taking into account sources of potential bias or imprecision. Discuss both direction and magnitude of any potential bias<br>*Limitations are extensively discussed in the first part of the Discussion.* |
| Interpretation | 20 | ☒Give a cautious overall interpretation of results considering objectives, limitations, multiplicity of analyses, results from similar studies, and other relevant evidence<br>*We extensively discuss all the limits in the interpretation of our study findings through the Discussion section* |
| Generalisability | 21 | ☒Discuss the generalisability (external validity) of the study results<br>*Generalizability of study findings is discussed* |
| **Other information** | | |
| Funding | 22 | ☒Give the source of funding and the role of the funders for the present study and, if applicable, for the original study on which the present article is based<br>*Source of funding and role of funders are given.* |

*Give information separately for exposed and unexposed groups.

**Note:** An Explanation and Elaboration article discusses each checklist item and gives methodological background and published examples of transparent reporting. The STROBE checklist is best used in conjunction with this article (freely available on the Web sites of PLoS Medicine at http://www.plosmedicine.org/, Annals of Internal Medicine at http://www.annals.org/, and Epidemiology at http://www.epidem.com/). Information on the STROBE Initiative is available at www.strobe-statement.org.
